# Platelet dysfunction reversal with cold-stored vs. room temperature-stored platelet transfusions

**DOI:** 10.1101/2023.09.17.23295666

**Authors:** Valery J. Kogler, Jeffrey A. Miles, Tahsin Özpolat, S. Lawrence Bailey, Daire A. Byrne, Morgan Bawcom-Randall, Yi Wang, Hannah J. Larsen, Franklin Reed, Xiaoyun Fu, Moritz Stolla

**Author notes:** Both authors contributed equally (Valery J. Kogler, Jeffrey A. Miles). Corresponding author: Moritz Stolla, M.D. Bloodworks Northwest Research Institute 1551 Eastlake Ave, Suite 100, Seattle, WA 98102 •. Source to support this project: M.S. received funding from the NIH (1R01HL153072-01), Department of Defense, W81XWH-12-1-0441, EDMS 5570. Registration: NCT03787927. Dr. Stolla received research funding from Terumo BCT and Cerus Corp. All other authors have no conflict of interest to disclose.

## Abstract

**Background:** Platelets are stored at room temperature for 5-7 days (RSP). Due to frequent and severe shortages, the FDA recently approved up to 14-day cold-stored platelets in plasma (CSP). However, the post-transfusion function of CSP is unknown and it is unclear which donors are best suited to provide either RSP and/or CSP.

**Objective:** To evaluate the *post-transfusion* function and predictors of *post-transfusion* function for platelets stored for the maximum approved storage times (7-day RSP, 14-day CSP) in healthy volunteers on acetylsalicylic acid (ASA).

**Methods:** We conducted a randomized cross-over study in ten healthy humans. Subjects donated one platelet unit stored at either RT (RSP) or 4 °C (CSP) based on randomization. Before transfusion, subjects ingested ASA to inhibit endogenous platelets. Transfusion recipients were tested for platelet function and lipid mediators. Platelet units were tested for lipid mediators only. A second round with transfusion of the alternative product and an identical testing sequence followed.

**Results:** RSP reversed platelet inhibition significantly better in αIIbβ3 integrin activation-dependent assays. In contrast, CSP led to significantly more thrombin generation in recipients, which was not dependent on platelet microparticles, but CSP themselves. Lysophosphatidylcholine-O (Lyso-Platelet Activating Factor) species levels predicted the procoagulant capacity of CSP. In contrast, polyunsaturated fatty acid concentration predicted the aggregation response of RSP.

**Conclusion:** We provide the first efficacy data of extended-stored CSP in plasma. Our results suggest that identifying ideal RSP and CSP donors is possible and pave the way for larger studies in the future.

**Graphical Abstract 1:**
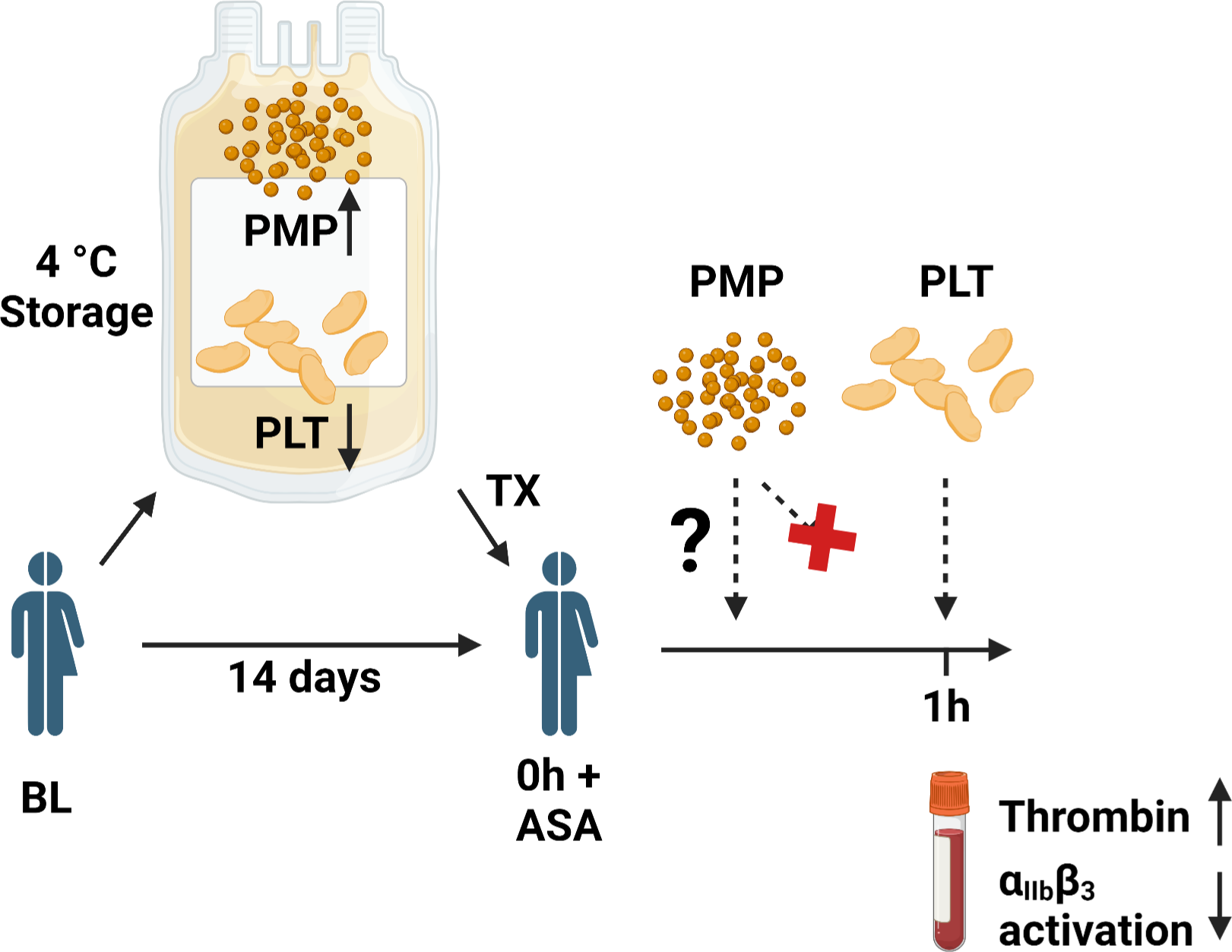
Overview of CSP function after 14 days of storage (Created with Biorender)

## Introduction

Platelets are stored for up to 5-7 days at room temperature and are transfused to bleeding patients and those at risk of bleeding. The factors that determine platelet function upon transfusion are unknown. While some platelet storage markers correlate with circulation time and recovery, stored platelet *post-transfusion function* is currently impossible to predict *pre-transfusion* by *in vitro* tests. One parameter that recently regained attention is storage temperature. Room temperature storage of platelets is the current gold standard because it maximizes circulation time. However, maintaining room temperature-stored platelets (RSP) is difficult and expensive; they are the most common cause of transfusion-transmitted infection, and recent clinical trials cast doubt on their efficacy and safety.^1,2^ Cold platelet storage (CSP) could alleviate these shortcomings.^3^ However, the most promising data on the efficacy of extended stored CSP comes from *in vitro* testing showing a procoagulant phenotype combined with superior or similar integrin function as RSP^.3–8^ During the COVID-19 pandemic, the FDA approved a variance of CSP stored for up to 14 days to reduce platelet shortages. Recently, the need for a variance was relinquished, effectively allowing all blood banks in the US to manufacture and transfuse up to 14-day-stored CSP (“if RSP are unavailable or not practical”), despite a paucity of post-transfusion data.^9^ A recent pilot trial compared extended-stored CSP to a historic RSP control group in cardiac surgery patients. In that study, RSP and CSP were stored in additive solution (platelet additive solution-E [PAS-E], SSP+, Macopharma, Tourcoing, France), a platelet preparation not licensed in the US^.10^ A recent retrospective study found delayed cold-stored CSP (i.e., those that were first RT-stored, then cold-stored just before expiration) associated with increased postoperative transfusion requirements.^11^ Previous reports on the post-transfusion function of CSP yielded contradictory results, but the relevance of these reports is hampered by short storage times, outdated manufacturing methodologies, the use of the bleeding time as a functional read-out, low sample size, and data corrections.^12–17^ While the body of literature supporting the advantageous *in vitro* function of extended-stored CSP is expansive, it is currently unknown to what extent, if any, the *in vitro* function of CSP translates into *in vivo* or post-transfusion function.

Adding urgency to the subject is the increasing difficulty of maintaining an adequate platelet inventory for blood centers and hospitals with current storage conditions, especially during the COVID-19 pandemic.^18–20^ The potential availability of a platelet product with a comparable hemostatic potential and extended shelf life is particularly advantageous for austere medical settings. Therefore, there is an urgent need for *in vivo* and post-transfusion human data to inform the current and future use of CSP and future regulatory assessments of this product.

Therefore, in the current study, we sought to address two critical knowledge gaps: 1) We compared the post-transfusion function of 7-day RSP (the current clinical maximum) with 14-day plasma-stored CSP (the recently approved maximum allowable storage time per FDA) in autologous, acetylsalicylic acid (ASA)-treated healthy humans, and 2) we tested for oxylipins, lysoglycerophospholipids (LPLs), and polyunsaturated fatty acids (PUFAs) in storage bags and recipients to identify predictors for post-transfusion function in recipients of CSP and RSP.

## METHODS

### Healthy human subjects research (including CONSORT criteria)

We conducted a randomized crossover trial in healthy humans comparing 14-day CSPs to the standard-of-care 7-day RSPs. Volunteers were recruited via paper and internet ads from December 2020 to September 2021 and enrolled at the Bloodworks Northwest Research Institute in Seattle, WA. Inclusion criteria were self-reported good health, age between 18-59 years, temperature: ≤ 99.5°F, systolic blood pressure ≤ 180 mmHg; diastolic ≤100 mmHg, heart rate 40 to 100 BPM. Exclusion criteria were personal or family bleeding history, a history of thrombosis, a history of or currently prescribed antiplatelet or anticoagulant agents, or other drugs known to significantly affect platelet function (full list of criteria in the protocol, available in the supplement).

Participants were randomly assigned to receive CSPs or RSPs first. No formal blinding was performed. In each period, participants underwent plateletpheresis with storage at either room temperature or 4°C. Acetylsalicylic acid (ASA, 325mg) was administered 24h before transfusion. Testing was done at baseline, pre-transfusion (post-ASA), 1h, 4h, and 24 h after transfusion. The prespecified primary endpoint was platelet αIIbβ3 (GPIIb-IIIa) integrin activation (VerifyNow ASA, Accumetrics Corp) in Aspirin Reaction Units (ARU) at 1h post-transfusion to measure the reversal of ASA antiplatelet therapy by platelet transfusion.^5^ This assay measures αIIbβ3-mediated aggregation of platelets in response to arachidonic acid in whole blood (normal: > 550 ARU, values below indicate platelet dysfunction). Secondary endpoints included ARU at 4h and 24h. Adverse events were assessed at all study visits and by participant reports. A protocol change to include a no-transfusion control group was planned but not implemented. The institutional review board (wcg-IRB) approved the study. Written informed consent was obtained. We conducted the study following the Declaration of Helsinki. All authors had access to the reported data.

### VerifyNow ASA

VerifyNOW for ASA (Accumetrics Corp, San Diego, CA) was a sent-out test to the University of Washington clinical laboratory at Harborview (Seattle, WA). Briefly, whole blood was collected in a 3.2% sodium citrate tube, transported to the lab, and inserted into the VerifyNOW instrument. The platelet aggregation induced by arachidonic acid (AA) is measured as Aspirin Reaction Units (ARU), a read-out of light transmission through the sample. Normal ARUs are higher than 550. An ARU <500 indicates platelet dysfunction, e.g. by cyclooxygenase-1 inhibition with ASA.

### Statistics

Based on previous study results, a minimum sample size of 6 was required to provide >98% power to detect a difference of 130 ARU for the primary endpoint between CSPs and RSPs. The protocol specified analysis plan was for a 2-tailed, paired t-test was used for statistical analysis. Because two volunteers only completed one treatment period (1 CSP and 1 RSP), an unpaired posthoc analysis was also performed. A p-value of ≤ 0.05 was considered significant (GraphPad Software, Prism 6.05). Pearson’s Correlation Coefficient was calculated to test for linear correlation. LC-MS/MS-MRM data was analyzed with the help of MetaboAnalyst Software 5.0.^21^

## RESULTS

### Healthy human demographics and recruitment

Over ten months, ten healthy participants were enrolled. One participant was excluded because of abnormal baseline platelet function. One subject did not complete the RSP period due to the low pH of the stored platelets and one subject did not complete the CSP period due to a scheduling conflict (Supplemental Figure 1), but both completed the corresponding alternative transfusion arm). In aggregate, nine data sets are available for both treatment periods, and eight of these represent matched (paired) data, while two additional subjects provided an unpaired data set. Participant baseline characteristics are summarized in Supplemental Table 1.

### Platelet unit characteristics and safety assessment

All platelet units passed quality control assessments after storage except for one RSP which had to be discarded due to low pH. All transfused platelets were tolerated well by the recipients, and one adverse event occurred (headache) that was considered unrelated to transfusion because it occurred 48 hours after the invention. The platelet concentration in the storage bag did not differ at baseline (Supplemental Figure 2A). Over storage time, cold temperature reduced the platelet concentration significantly compared to RT (Supplemental Figure 2A) presumably due to microaggregates.^22^ However, the absolute number of transfused platelets and the number of transfused platelets as a percentage of absolute (endogenous and transfused) circulating platelets did not significantly differ between the two treatment groups (Figure 1B, Supplemental Figure 2B)), even though we did not adjust the platelet concentration before transfusion.

**Figure 1.**
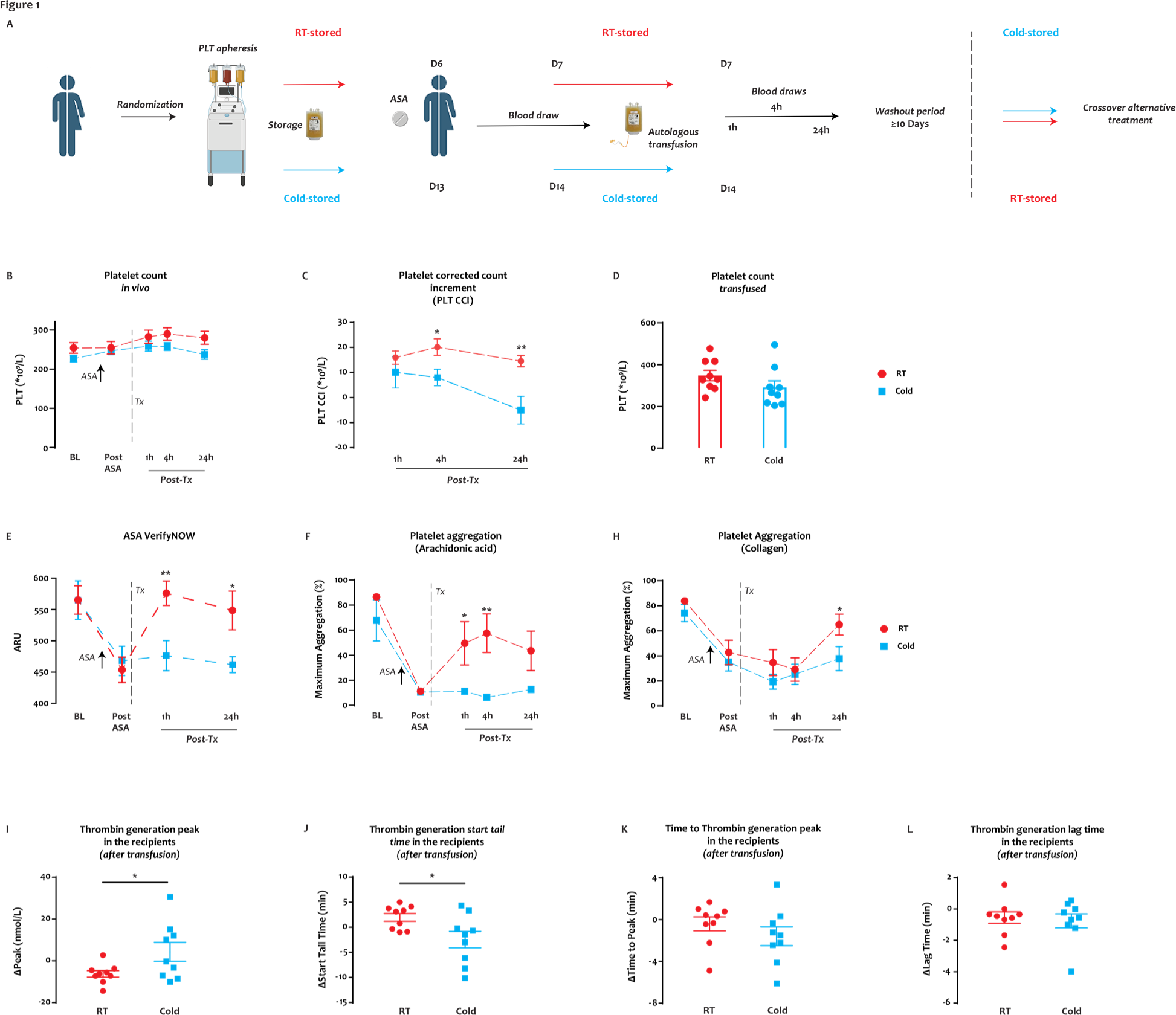
Effect of RSP and extended-stored CSP transfusion in healthy humans on aspirin. **(A)** In each of the two periods, healthy humans underwent plateletpheresis with platelet storage (randomized to RT or 4°C), ASA loading 24 hours prior to transfusion, and autologous platelet transfusion with blood assessments at baseline (BL), pre-transfusion (ASA), and at multiple time points after transfusion (1h, 4h, 24h). Subjects completed an RSP period (receiving 7-day RT platelets) and a CSP period (receiving 14-day 4°C platelets) as outlined in this randomized, crossover study overview schematic. **(B)** The platelet count and **(C)** corrected count increment (CCI) during each period (red circles – RSP period; blue circles – CSP period; 4h *p=0.0204, 24h *p=0.0047). **(D)** The number of RT (red circles) and 4°C (blue circles) total platelets transfused. **(E)** Platelet arachidonic acid-stimulated αIIbβ3 activation was measured by Aspirin VerifyNow (1h **p=0.0055, 24h *p=0.0198). **(F)** Maximum aggregation in response to 1mM arachidonic acid (1h *p=0.0347, 4h **p=0.0081). **(G)** Maximum collagen (2.5 ug/mL) aggregation was also measured at all time points (24h *p=0.0493). The thrombin generation potential of the subject’s whole blood-derived platelet-rich plasma was measured immediately before and 1h after transfusion. Results are reported as the change from pre-transfusion to 1h post-transfusion value for **(H)** thrombin generation peak (*p=0.0429), **(I)** start tail time (*p=0.0266), **(J)** time to thrombin peak, and **(K)** lag time. Data are shown as mean ± SEM, (H-K shown with individual values). Unpaired data, n=7-9, individual p values are shown in the text above.

### Platelet transfusion parameters

There was no significant difference in absolute recipient platelet counts before and after transfusion between RSP and CSP groups (Figure 1C), but when taking the recipient body surface area, the number of transfused platelets, and the platelet count increment into account (corrected count increments, CCI), we found significantly lower CCIs in the cold-stored platelet transfusion arm after 4 hours and 24 hours (Figure 1D). The average time between the two periods was 22 days, and the shortest was 11 days (Supplemental Table 1).

### Function of transfused platelets

We tested for platelet function by VerifyNow ASA, a clinically used assay based on arachidonic acid-mediated platelet αIIbβ_3_ integrin activation in whole blood. We observed normal platelet function at baseline and appropriate inhibition of platelet function after ASA dosing (Figure 1E). Transfusion of RSP significantly improved platelet function by VerifyNow ASA one hour (which was the prespecified primary endpoint of the study) and 24 hours (prespecified secondary outcome) after transfusion compared to CSP (p=0.0055 and p=0.0198, respectively) (Table 1, Figure 1E). Transfusion of CSP failed to overcome the effect of ASA at any time. Similarly, platelet aggregation in response to arachidonic acid showed improved platelet response 1 hour (p=0.03) and 4 hours (p=0.008) after transfusion with RSP compared to CSP (Figure 1F). ASA led to a modest inhibition of collagen-mediated platelet aggregation, and recipients of RSP were significantly more responsive to collagen only 24 hours after transfusion (Figure 1G).

**Table 1:** Primary and secondary outcomes.

|  | Unpaired analysis |  |  |  | Paired analysis |  |  |  |
| --- | --- | --- | --- | --- | --- | --- | --- | --- |
|  | CSP | RSP | Difference | P value | CSP | RSP | Difference | P value |
|  | n = 9 | n = 9 |  |  | n = 8 | n = 8 |  |  |
| Primary Outcome |  |  |  |  |  |  |  |  |
| ASA VerifyNOW 1h, mean ARU (95% CI) | 472 (420, 525) | 576 (531, 621) | -104 (-168, -40) | 0.003 | 463 (407, 519) | 569 (520, 618) | -106 (-174, -38) | 0.02 |
|  | Unpaired analysis |  |  |  | Paired analysis |  |  |  |
|  | CSP | RSP | Difference | P value | CSP | RSP | Difference | P value |
|  | n = 9 | n = 9 |  |  | n = 8 | n = 8 |  |  |
| Secondary Outcome |  |  |  |  |  |  |  |  |
| ASA VerifyNOW 24h, mean ARU (95% CI) | 462 (433, 491) | 549 (477, 620) | -87 (-158, -16) | 0.02 | 462 (428, 496) | 538 (460, 616) | -76 (-153, 1) | 0.06 |

Numerous *in vitro* studies have shown a pro-coagulant phenotype of CSP units with a potential contribution from phosphatidylserine-positive platelets and platelet-derived microparticles (PMP) released during storage.^8,23–25^ It has never been investigated whether CSP can alter thrombin generation characteristics after transfusion in humans. To clarify the potential procoagulant role of CSP transfusion, we measured thrombin generation before and 1 hour after transfusion in PRP derived from recipients’ whole blood. We found a statistically significant increase in peak thrombin generation in recipients after CSP transfusion compared to RSP (p=0.04) (Figure 1H). In accordance, we found a trend for a shorter time to peak thrombin generation (Figure 1I) and a significantly shorter tail time (Figure 1J). No significant change was seen for lag time (Figure 1K).

### Platelet Microparticles (PMP) before and after transfusion

To test whether cold-stored platelets themselves or platelet-derived microparticles from 4 °C units were responsible for the increase in peak thrombin, we first assessed the platelet-poor plasma (PPP) of stored platelet units for PMP (Figure 2A). To allow for the detection of small PMP to as low as 100 nm, we utilized a CytoFLEX flow cytometer and validated small PMP detection using a multicolor approach, which allowed simultaneous assessment of CD41a, lactadherin, and CD62P (Figure 2A-F). Small microparticles were significantly more abundant in CSP than RSP with all gating strategies, pan-platelet marker positive events (CD41a) (Figure 2G), phosphatidyl serine (PS) positive events (Figure 2H), and P-selectin positive events (Figure 2I), while large PMPs were only significantly more abundant in CSP than RSP in the pan-platelet gate. Similarly, within CSP and RSP small PMP were significantly more abundant than large PMP (Figure 2G-I), except for P-selectin-positive PMP in RSP. Less than 1% of CSP PMP were large, suggesting that previous reports on CSP PMP may have markedly underestimated the total number of PMP in stored units.^7,26,27^ Smaller PMP carried significantly more PS on their surface (Figure 2H, and significantly more P-selectin (Figure 2I), increasing procoagulant effect and interaction with white blood cells, along with other potential sequelae.^28^ We then tested stored PPP with a thrombin generation assay and observed a significantly increased peak thrombin generation and a reduced tail time from CSP units (Figure 2G, H). Also, we observed trends for shortened time to peak and shortened lag times with pre-transfusion PPP (Figure 2 I, J).

**Figure 2.**
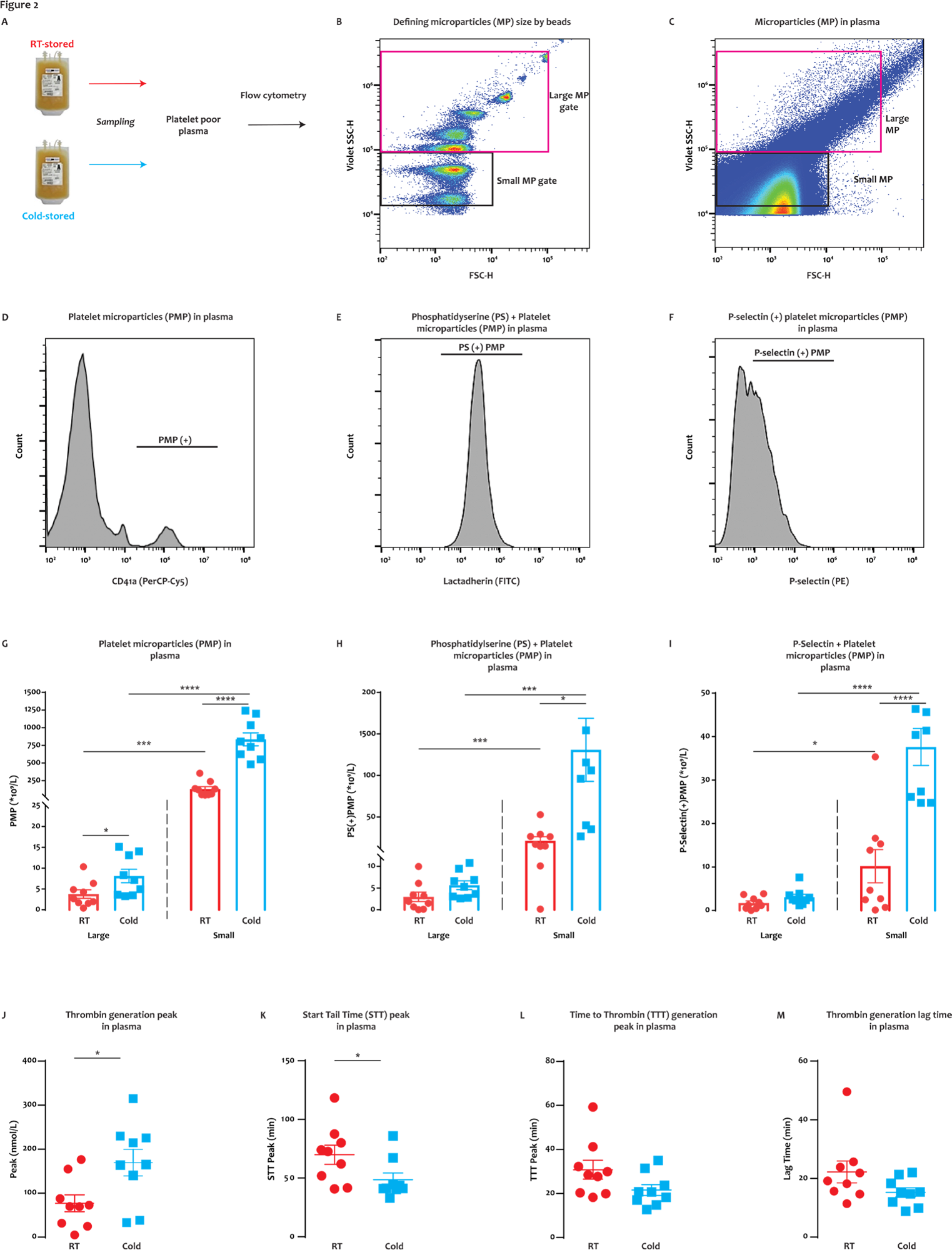
Increased small platelet microparticles and thrombin generation with 4°C stored platelets *in vitro*. **(A)** After platelet storage at both RT and 4°C conditions, platelet-poor plasma was prepared by centrifugation with one freeze-thaw cycle prior to testing. **(B)** Utilizing the Cytoflex flow cytometer, and GigaMix flourescent beads (various sizes from 0.1 µm to 0.9 µm), we identified MP with a large MP size gate (1 µm-eq. to 0.16 µm-eq.) and a small MP size gate (between 0.16 µm-eq. to 0.1 µm beads). **(C)** The gates were applied to microparticles in plasma. **(D)** Within the respective large and small MP size gates, platelet MPs (PMPs) were defined as all CD41a+ events. Both large and small PMP were further gated for lactadherin **(E)** or CD62P **(F)** positivity within the CD41+ gate. **(G)** Platelet microparticles (PMP) of both RT and 4 °C platelets were identified as events within small and large size gates that were CD41a+ and the concentration of each reported (large *p=0.0369, small ****p<0.0001). **(H)** The concentration of large and small PMP that were also positive for lactadherin (small *p=0.0113) or **(I)** CD62P (small ***p=0.0002). The microparticle-mediated thrombin generation potential of PPP from storage bags was measured and reported as **(J)** thrombin generation peak (*p=0.0202), **(K)** lag time, **(L)** time to thrombin peak, and **(M)** start tail time (*p=0.0479). Data are shown as mean ± SEM and individual values. Unpaired data, n=9, individual p values are shown in the text above.

To clarify the possible post-transfusion relevance of the additional PMP with CSP, we assessed the number of large and small PMP before and after transfusion in recipients (Supplemental Figure 2A). We did not observe any increase in large or small PMP, not by size, PS, or P-selectin (Supplemental Figure 2B-D) after transfusion of either platelet product, suggesting clearance of the PMP occurred within the first hour. Accordingly, we did not find more thrombin generation in PPP obtained after CSP transfusion (Supplemental Figure 2E-H). In fact, PPP derived from whole blood 1 hour after RT-stored platelet transfusion led to significantly more thrombin generation than after CSP transfusion (Supplemental Figure 2E). Together, these findings suggest our observed increase in thrombin generation one hour after CSP transfusion (Figure 1E) was dependent on phosphatidyl serine (PS) expressed on transfused platelets and independent of PMP.

### Lipid metabolites of ASA-treated individuals before and after drug administration

To improve our understanding of platelet lipid changes after transfusion in healthy humans on ASA, we first measured the changes after a single loading dose of ASA. We tested for oxylipins, PUFAs, and lysoglycerophospholipids, such as lysophosphatidylcholine (LPC) species, lysophosphatidylethanolamine (LPE), lysophosphatidylserine species (LPS), and lysophosphatidylinositol species (LPI). As expected, we found significantly lower COX-1-dependent oxylipins, such as thromboxane B2 (TxB2) 12-HHTrE, and 11-HETE (Supplemental Table 2) in ASA-treated volunteers compared to baseline. Thromboxane B2 levels trended lower in ASA-treated individuals (p=0.06, Supplemental Table 2). In contrast, certain LPE, LPS, LPI species were significantly higher after ASA administration (Supplemental Table 2). In accordance with a previous report, we did not find significant differences before and after ASA administration in LPC species.^29^

### Lipid metabolites in stored platelets

We observed a heterogeneous storage response for oxylipins and PUFAs (Figure 3 A); however, LPC species increased more broadly and significantly in RSP than CSP (Figure 3 A,B,D). LPS species increased at both storage temperatures, but more uniformly in RSP (Figure 3 A, B). Changes in LPE species were not uniform with some species increasing and some decreasing, however, the decrease of some LPEs was more pronounced in RSP than CSP (Figure 3B, E). Oxylipins, such as 12-HETE, increased to a similar degree at both storage temperatures, but the number of oxylipins with significance and more change from baseline was higher in CSP (Figure B, E) In a principal component analysis (Figure 3 C,E) we found stored CSP samples mixed with baseline values in the 2D scores plot (PC1 versus PC2), while stored RT platelets were clearly separated from baseline (Figure 3, C,F) Principal component 1 explained 27 % of the variance between baseline and stored CSP and RSP samples (Supplemental Figure 4 A). The top 15 metabolites from the variable importance in projection plot featured LPC species (including LPC-O and LPC-P species), LPE, and LPI species (Supplemental Figure 4 B). The concentrations of total LPCs were significantly higher in RSP compared to CSP, including all LPC-O (16:0, 18:0, 18:1, 18:2) species (also known as lyso-platelet activating factor species) (Figure 4 A, B). There was a trend for lower total LPEs in CSP compared to RSP, but this did not reach statistical significance, possibly because some isolated LPEs decreased more in RSP than in CSP (Figure 3 B, E). Certain LPS species were significantly higher in CSP (Figure 4 E), and some LPI species were higher in RSP. Because of the high donor-to-donor variability, we also compared the relative increase normalized to baseline (Supplemental Figure 5). Total LPC, LPE, and LPI levels increased more in RSP than CSP, while total LPS species increased significantly more in CSP. Interestingly all LPC-O species (also known as Lyso-platelet activating factor [LPAF] species) increased significantly more in RSP than CSP (Supplemental Figure 5 E-H). The linoleic acid-derived oxylipin 12,13-diHOME was the only oxylipin that differed significantly between CSP and RSP when normalized to baseline, while thromboxane B2, 12,13 EpOME and 5-HETE approached statistical significance (Supplemental Figure 5, I-L).

**Figure 3:**
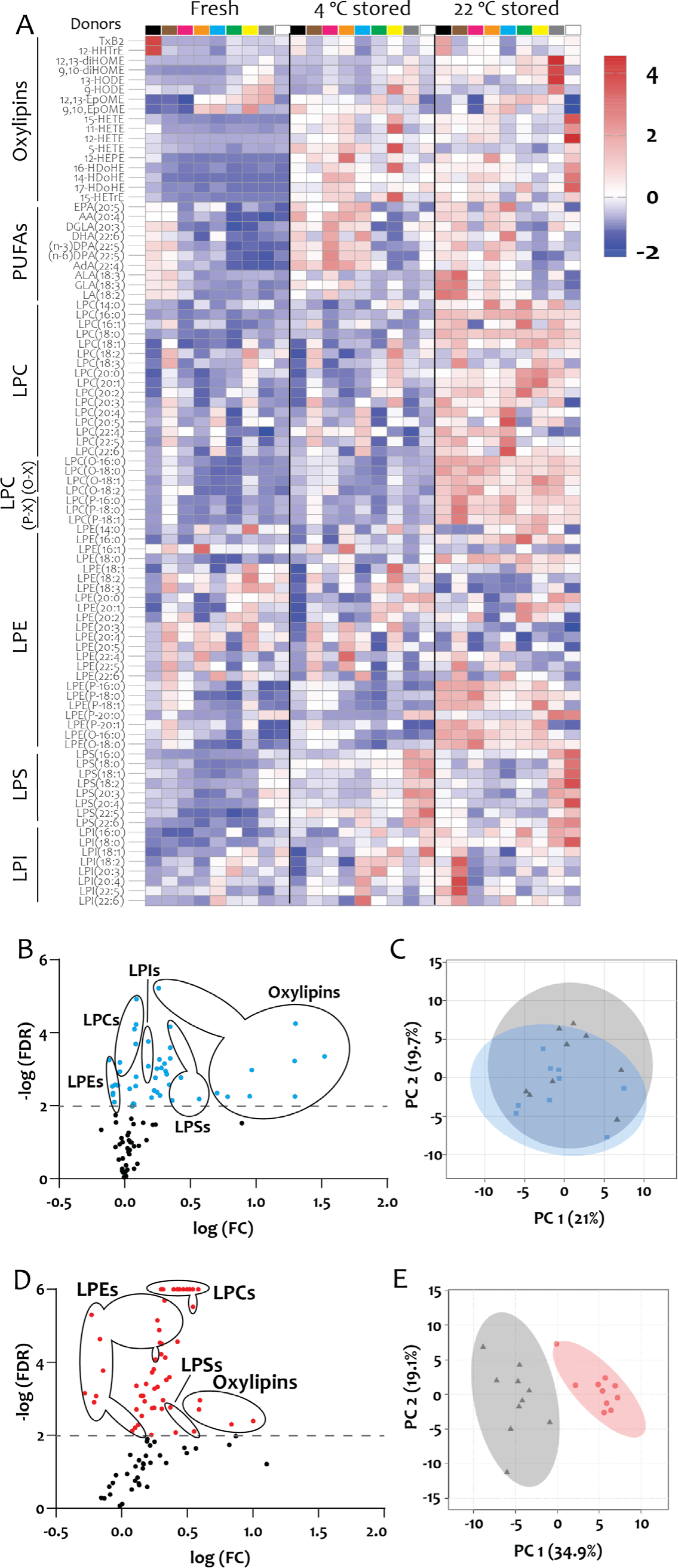
Bioactive lipid mediators in CSP and RSP storage bags. Data were mean-centered and divided by the standard deviation of each variable to normalize. **(A)** Heat map with individual lipid mediator levels at baseline (Fresh) and after 14 days of 4 °C storage, and 7 days of RT storage. Color codes for individual donors in first row. The baseline (fresh) data was averaged from two diffrerent collections. **(B)** Volcano plots of CSP, plotted as logarithmic false discovery rate (Q=1%), and logarithmic fold change, and **(C)** 2D scores plot of principal component analysis of CSP **(D)** Volcano plots of RSP, and **(E)** 2D scores plot of principal component analysis of RSP. N=9 (only paired data), Statistical significance was assessed with a false discovery rate (FDR) of Q = 1% (89 comparisons).

**Figure 4:**
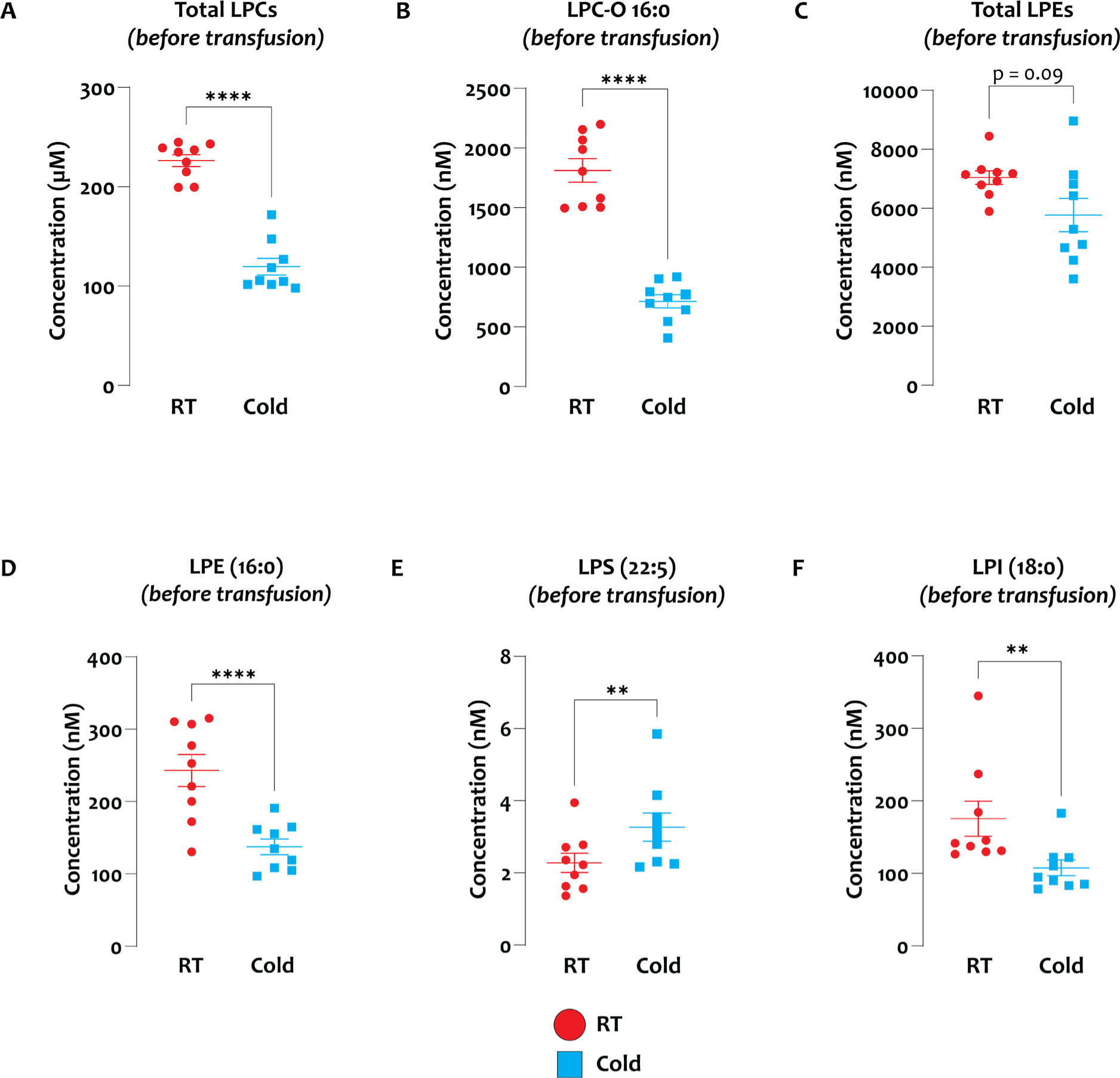
Lipid mediator concentration comparison between RSP and CSP. **(A-F)** Samples at the end of storage were compared between RSP and CSP. Data shown as individual values with mean and SEM. N=9 (Paired Analysis) with α = 0.05 for total LPGL comparisons with Holm Sidak correction for multiple (4) comparisons. Individual species were tested with false discovery rate of Q=1% (89 comparisons).

### Lipid metabolites of transfusion recipients

We normalized the recipient lipid mediator data to post ASA administration to detect specific differences between transfusion of CSP and RSP in recipients. In recipients, total LPC, and LPI levels were significantly higher only 24 hours after transfusion of RSP, while LPE species were significantly higher at 1h, 4h, and 24h after transfusion of RSP compared to CSP (Figure 5 A, B-D). Linoleic acid oxylipins, derived from CYP and lipoxygenases, such as diHOMEs, EpOMEs, and HODEs were significantly higher or approached significance in the recipients of RSP than CSPs (Figure 5 E-J).

**Figure 5:**
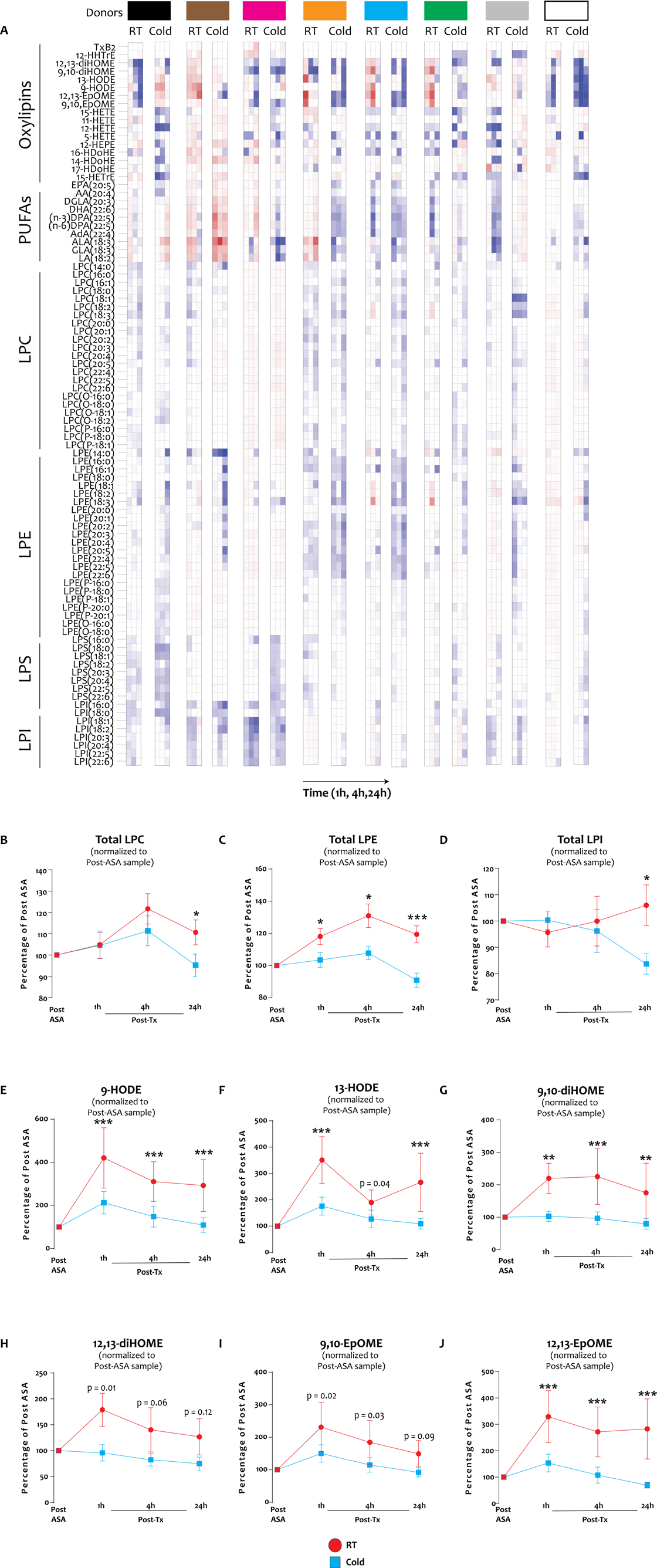
Bioactive lipid mediators in CSP and RSP transfusion recipients. **(A)** Heatmaps of individual transfusion donor/recipients (color-codes - first row) normalized to post-ASA (pre-transfusion). Individual 3 squares show values for 1h, 4h, and 24h after transfusion. (**B-M**) Individual and pooled (“total”) bioactive lipid mediators normalized to post-ASA (pre-transfusion). Individual lipid markers were tested with two-tailed, paired t-test with Q=1% FDR (89 comparisons), and total (sum) lipids with α = 0.05 and Holm-Sidak method for multiple (4) comparisons. Data are shown as mean ± SEM, n=8 (only paired recipient data included), *p≤0.05, **p<0.01, ***p<0.001.

### Storage and recipient lipid metabolites predictors of post-transfusion platelet function

To identify predictors of post-transfusion platelet function we performed correlation analyses of our targeted lipid panel (both storage unit and recipient) and the corresponding platelet function data (from recipients). This approach allowed us to identify markers in the storage unit and the recipient that would predict post-transfusion performance of stored platelets. Linoleic acid-derived oxylipins (HODES) in storage bags predicted collagen-induced platelet aggregation in CSP recipients (Figure 6 A-B).

**Figure 6:**
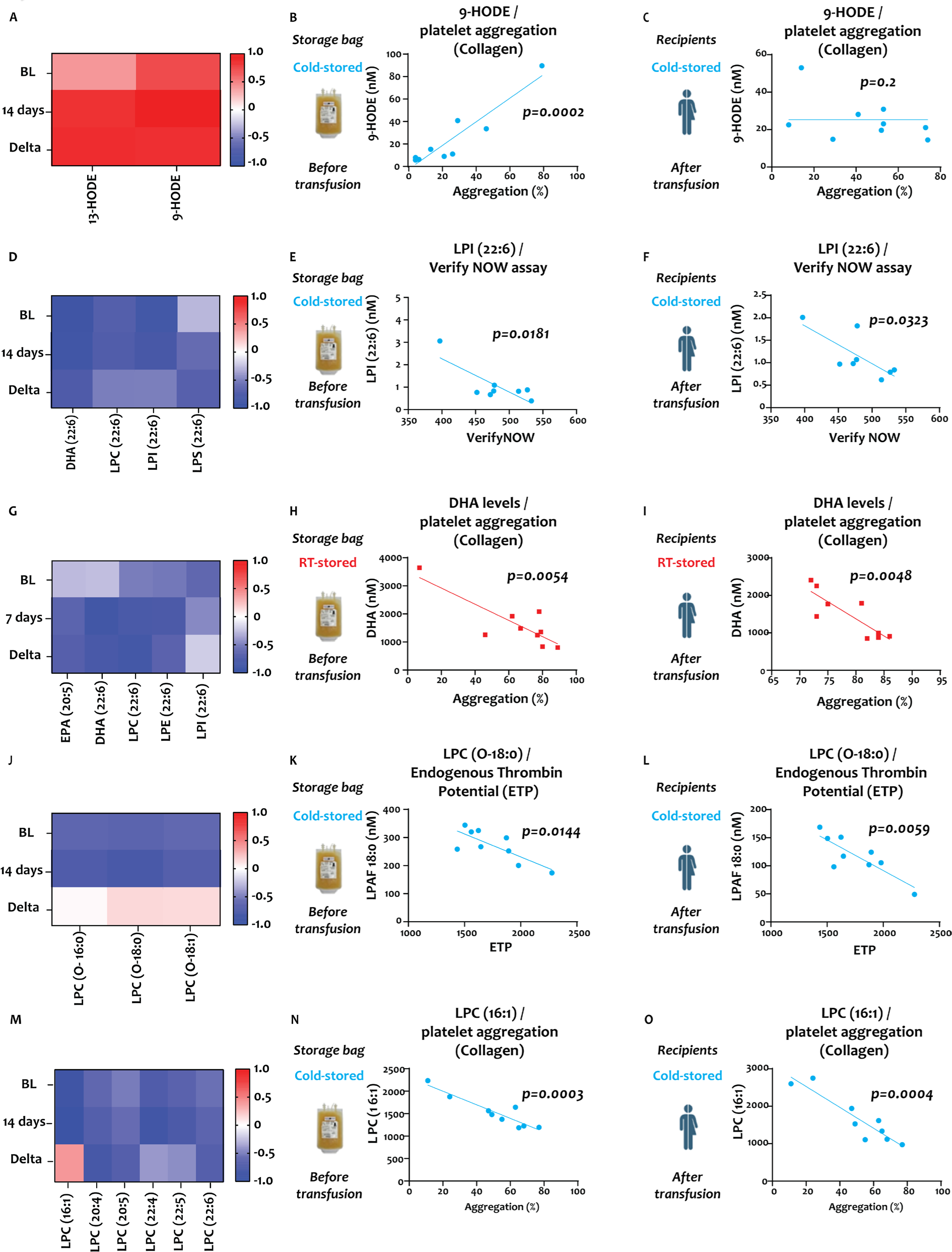
Correlation of lipid mediators and platelet function parameters. Left column (**A, D, G, J, M**) shows r-value heat maps for baseline (BL), end of storage (7 and 14 days, respectively), and for the difference between BL and end of storage (Delta). Middle column (**B, E, H, K, N**) shows the correlation between platelet functional parameters and storage bag lipid mediator concentration. The right column (**C, F, I, L, O**) shows the correlation between platelet functional parameters and recipient bioactive lipid mediator concentration. Data are shown as heatmaps and linear regression analysis plots. N=8 in each group, paired data only, and individual p values are shown in each figure.

However, the correlation was no longer detectable upon transfusion in the recipient (Figure 6 C). LPI species from CSP units and CSP recipients correlated negatively with αIIbβ_3_ integrin platelet function in whole blood (Figure 6 D-F). PUFAs, including those conjugated to lysoglycerophospholipids correlated negatively with platelet aggregation in plasma (Figure 6 G-I). As shown above, CSP’s main function appeared to be to provide a procoagulant surface after 14 days of storage. Therefore, a predictor of thrombin generation was of high interest. Indeed, we found LPC-O (LPAF) species to negatively correlate with endogenous thrombin generation potential (ETP) of stored CSP platelets and platelets from CSP transfusion recipients (Figure 6 J-L). RSP were trending toward a significance for a correlation between LPC-O species in the storage unit and ETP, however, no correlation was observed upon transfusion (Supplemental Figure 6 A-C). Because blood banks will have to assign the platelet storage temperature within the first hours after donation, we also tested for pre-storage correlation of LPC-O species. Indeed, pre-storage levels of LPC-O also predicted recipient ETP (Supplemental Figure 6 D). For CSP we also observed a negative correlation of LPC species with collagen-induced aggregation in both CSP storage unit and CSP transfusion recipient (Figure 6 S-U).

## DISCUSSION

A large body of pre-transfusion *in vitro* data suggest a superior function of CSP compared to RSP. Our study provides the first post-transfusion head-to-head comparison of CSP and RSP stored in plasma up to the maximum FDA-approved storage times. We evaluated the post-transfusion αIIbβ3 integrin function of CSP and RSP in recipient-derived platelet-rich plasma and whole blood. Thus far, this is the first study showing the reversibility of ASA with 7-day RSP (the maximum approved store time) in transfusion recipients. Previous studies showed that ASA can be overcome by adding RSP *ex vivo* or by transfusing frozen platelets to patients on ASA^.30,31^ Two studies from the 1970s suggested RSP could reverse the effect of ASA, but they used 3-day stored, pooled whole blood-derived platelets, and assessed efficacy by bleeding time only. These historic studies also used 4 °C platelet storage as a control. Their results showed that 4 °C storage led to a more rapid and potent reversal of the effect of ASA as measured by bleeding time, contradicting our findings.^15,17^ However, the difference between 3 and 14 days of storage is large enough to suggest that it could be responsible for the functional decrease of CSP observed in our study. In our previous cross-over study using aspirin and clopidogrel in healthy humans and comparing the effect of 5-day RSP and 5-day CSP we found equivalence in the VerifyNOW ASA assay, and light transmission aggregometry with arachidonic acid.^25^ While we still observed improvement with 7-day RSP in the current study, this effect was essentially non-existent with 14-day CSP contradicting a large body of *pre-transfusion* literature^.7,32,33^

In our study, CSP are significantly more pro-coagulant in transfusion recipients suggesting that the hemostatic effect of extended-stored CSP may be more akin to cryopreserved (i.e., frozen) or lyophilized platelets, albeit with a shorter circulation time^.34,35^ Surprisingly, a recent trial with lyophilized platelets did not show increased thrombin generation in transfusion recipients.^36^ This may have been due to the low dose tested in the trial. Yet, while we show a statistically significant increase in thrombin generation parameters with 4 °C platelet transfusion compared to control, it is unclear whether the difference we observed one hour after transfusion is clinically meaningful.

A historical assessment of 4 °C unit PMPs observed a trend for an increased number of smaller microparticles^.26^ To our knowledge, we provide the first data on CSP and RSP-derived PMP utilizing a high-resolution flow cytometer measuring PMP with a diameter as low as 100nm. In our study, PMP were not responsible for the increase in thrombin generation in recipients of CSP. Instead, we found transfused platelets to be responsible for the procoagulant effect. In contrast, PMP have been suggested to be critical for the hemostatic effect of cryopreserved (i.e. frozen) platelets^.37^ One of the limitations of our study is that our earliest testing time point was one hour after transfusion. Recipients of CSP may experience a more substantial, immediate but short-lived (< 1 hour) procoagulant impact after transfusion due to transfused PMP. Very limited data are available on the *in vivo* clearance kinetics of PMP from transfused platelets. A previous report in thrombocytopenic patients reports a half-life of large PMP of 5.8 hours using conventional flow cytometry^.38^ The clearance likely depends on the phenotype and size of PMP, as well as recipient factors. In contrast to other published data^38^, we did not see an increase in PMP, either after transfusion of RSP, or CSP. One possible explanation is that clearance of PMP is accelerated in healthy humans compared to multimorbid patients. Potential mechanisms include splenic sequestration, PMP-platelet, or PMP-endothelial adhesion. Another limitation of the study is that the PRP samples were analyzed fresh, whilst the PPP samples were frozen on the day of the experiment and analyzed in bulk, which could have affected the quality and procoagulant activity of the PMP in plasma. The relatively low inclusion number of ten healthy volunteers is a limitation, but by utilizing a cross-over design and thereby using each subject as their own control, we eliminated the between-subject variability. Cross-over designs also allow for achieving significance with fewer participants and experimenting with healthy humans per se limits variability compared to multimorbid patients. Furthermore, the number of participants in our study exceeded the number required to detect significant differences in the primary endpoint (1h ASAVerifyNOW) as per our power analysis (see statistical analysis).

Combining targeted lipid metabolite analysis of stored and transfused platelets before and after ASA dosing allowed us to not only analyze lipid levels but also to correlate them with functional data to identify predictors of RSP and CSP function, respectively. In a study with healthy humans on a 14-day low dose (81mg) ASA intervention, the authors also correlated collagen and arachidonic acid-induced aggregation of fresh platelets with oxylipin levels and found linoleic acid (LA)-derived oxylipins to be the only predictive markers.^39^ Likewise, we found LA-derived oxylipin HODE predictive of collagen-induced aggregation. LA is the most common dietary fatty acid, the LA-derived oxylipin 13-HODE prevents platelet adhesion to the vessel wall and inhibits platelet aggregation in rabbits^.40,41^ In contrast, we show that HODE levels during storage of CSP correlate positively with platelet aggregation. However, the doses used in the above publications were much higher than the levels we detected. PC-O-linked species (Platelet-activating factors [PAF]) are highly proinflammatory bioactive lipids released by various cells upon stimulation. We found LPC-O species (the deacetylated form of PC-O/PAF) to have a negative predictive effect on thrombin generation potential. LPC-O species inhibit platelet aggregation and thereby oppose the effect of PC-O species, but the negative correlation with thrombin generation after cold storage suggests that LPC-O species could also be inhibitors of their procoagulant function, which has not been described before^.42^ Importantly, low baseline (pre-storage) LPC-O levels were able to predict post-transfusion thrombin generation potential. We found thrombin generation to be the main function of extended-stored platelets in plasma. Therefore, in the future, measuring LPC-O levels may help to decide which platelets are most procoagulant and are best suited for cold storage. Of note, RSP did not show the same degree of correlation and significance suggesting that this finding is storage temperature-dependent. Numerous previous studies have described an inhibitory effect of ω-3 PUFAs on platelet aggregation, but the effect on stored platelet function and post-transfusion platelet function was unknown.^43–45^ In our study, PUFAs in the storage bag and recipients were strong predictors for reduced aggregation after transfusion. Whether this is predictive of decreased or increased protection from hemorrhage in transfusion recipients remains to be investigated. One previous study reported on the metabolomics of CSP but did not include bioactive lipid mediators or functional data.^46^

Our lipid mediator panel included a range of pro-inflammatory and anti-inflammatory mediators. Pro-inflammatory mediators were more prominent in RSP bags and recipients. For example, 12,13-diHOME was significantly higher in RSP than in CSP. Similarly, LPCs and LPIs, both pro-inflammatory, were significantly higher in RSP storage units and recipients. LPCs are known to inhibit platelet aggregation, secretion, and adhesion and could contribute to the surprising findings of recent clinical trials with RSP^.47–49^ LPIs cause vessel dilation^50^ and could thereby counteract hemostasis and mediate hypotensive transfusion reactions, although this needs further confirmation.

In contrast, anti-inflammatory and procoagulant mediators were more prominent in CSP. LPS species have been involved in the resolution of inflammation.^51^ Like its diacyl counterpart, phosphatidylserine (PS), Lyso-PS species also exert their procoagulant properties by supporting the assembly of the prothrombinase complex.^52^ CSP bags carried significantly more LPS species, and the total LPS increased more in CSP than in RSP when normalized to baseline. Accordingly, recipients generated significantly more thrombin after transfusion.

In summary, our study provides a first investigation of the post-transfusion function of RSP and CSP stored in plasma to their respective FDA-approved maximums. In our primary endpoint, we observed a significant increase in arachidonic acid-mediated integrin activation after RSP transfusion compared to CSP, while CSP improved thrombin generation characteristics one hour after transfusion. The clinical relevance of our data and surrogate outcomes in this healthy human crossover study is unclear but suggests limitations on the predictive value of many common *in vitro* assays for the post-transfusion function of stored platelets. While supporting our findings with mechanistic insight through targeted lipid mediator analysis and identifying markers with possible improved predictive value, caution is warranted until further data from randomized clinical trials in cancer, trauma or surgery patients are available.

### Authorship contributions

V.J.K and J.A.M. performed experiments, analyzed data, and wrote the manuscript. T.Ö. analyzed data, wrote the manuscript, and designed and created figures for the manuscript, S.L.B. and D.A.B. performed experiments and analyzed data. M.B. recruited and consented patients, and performed platelet collections, Y.W., H.J.L., F.R. performed experiments and analyzed data., X.F. designed experiments, and analyzed data, M.S. outlined the study, designed experiments, analyzed data, and wrote a first draft of the manuscript. All authors provided feedback on the manuscript and approved the final version of the manuscript.

## Data Availability

All data produced in the present study are available upon reasonable request to the authors

## Acknowledgments

M.S. received funding from the American Society of Hematology (ASH Scholar Award), NIH (1R01HL153072-01), and DoD (W81XWH-12-1-0441, EDMS 5570).

The authors would like to thank Renetta Stevens, Derek Nazareth, and Tena Petersen for administrative support. The authors would like to thank Amily Guo, Ph.D., and Xiaoping Wu Ph.D. for helping with the platelet microparticle analysis.

## Figure Legends

**Supplemental Figure 1:**
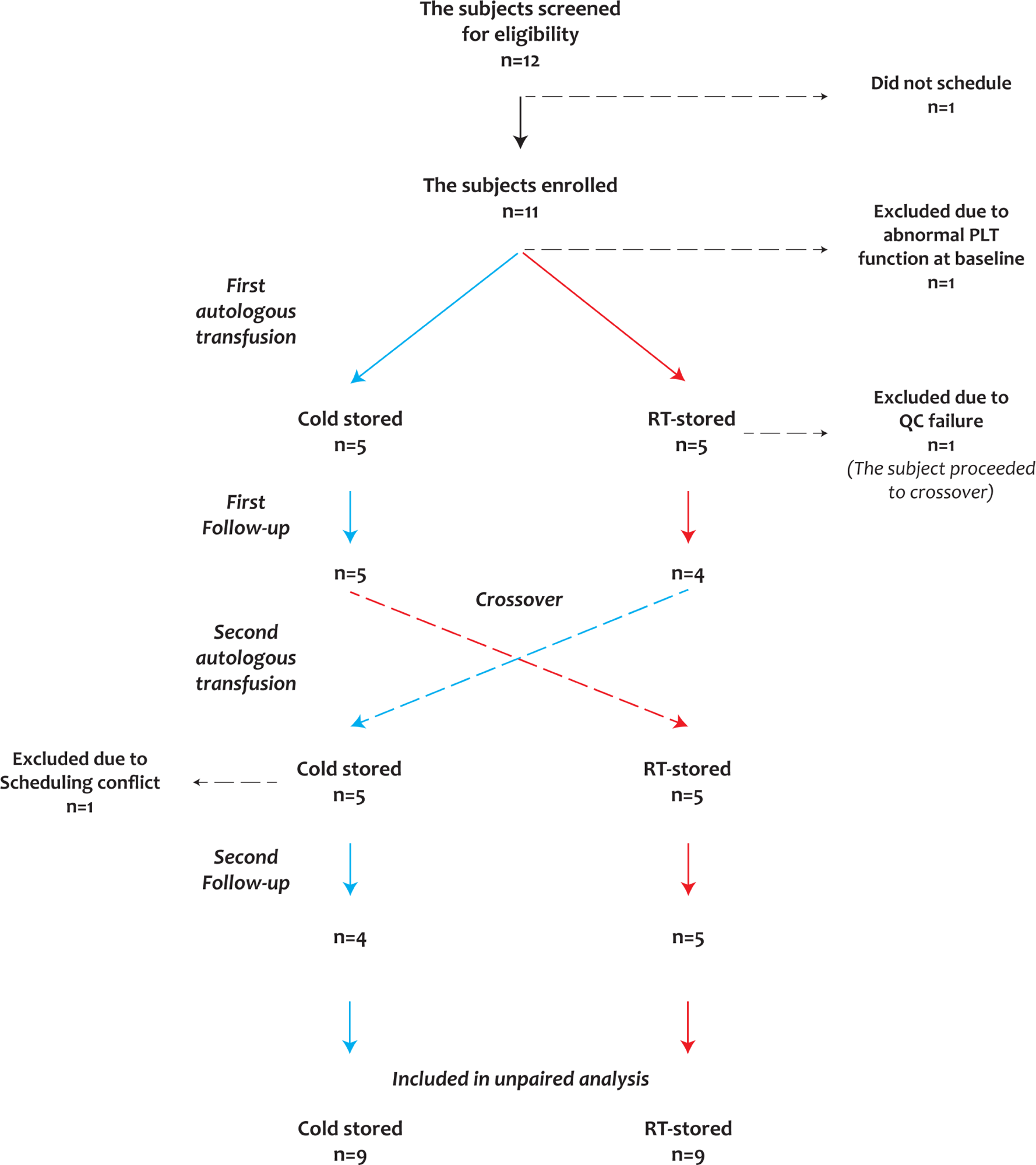
Flow chart of study outline.

**Supplemental Figure 2:**
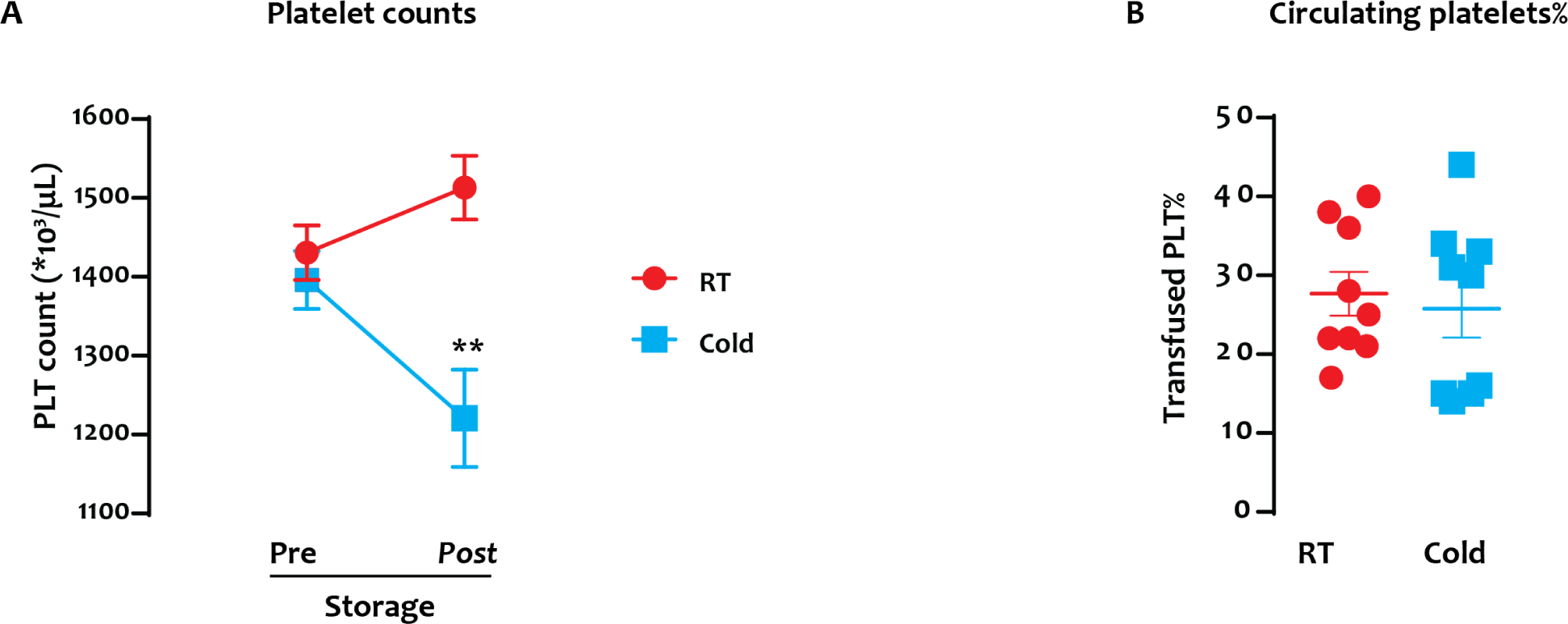
Platelet storage and post-transfusion numbers. **(A)** Platelet concentrations in the stored units were tested by ABX at the beginning and at the end of storage by hemocytometer. **(B)** The theoretical percentage of transfused platelets 1 hr after transfusion to total circulation platelets are shown as a percentage of all circulating platelets (Transfused PLT% = Transfused PLT yield / (Pre-transfusion total circulating platelets [calculated using standard CCI method] + transfused PLT yield) x 100). Data are shown as mean ± SEM (and individual values in B), N=9.

**Supplemental Figure 3.**
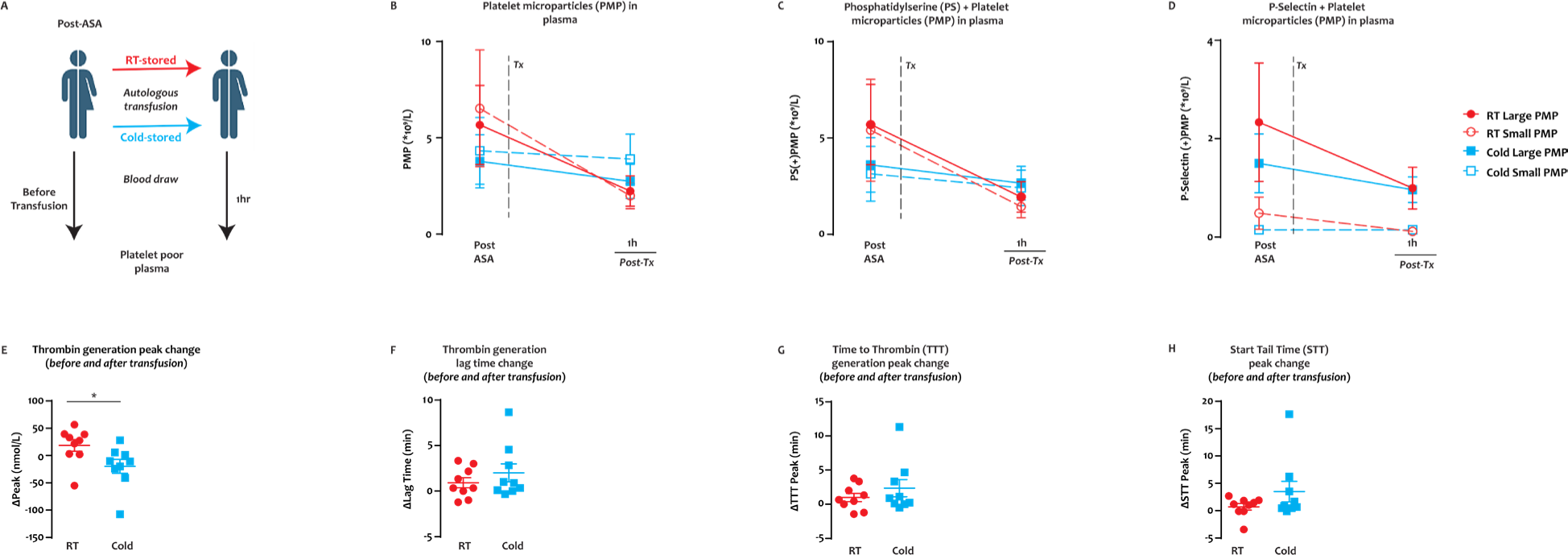
Detection of platelet microparticles and post-transfusion microparticle-mediated thrombin generation. **(A)** Whole blood samples were collected before and 1h after platelet transfusion. PPP was prepared and testing completed after one freeze-thaw cycle. **(B-D)** The concentration of large and small PMP for RSP (solid large red circles– large PMP, open red circles – small PMP) and CSP (solid blue squares – large PMP, open blue square – small PMP). No statistically significant differences were detected between treatment groups when **(B)** all PMPs (CD41+) were included, or when the concentration of **(C)** lactadherin and **(D)** CD62P+ PMP for both treatments were analyzed. The microparticle-mediated thrombin generation potential of subjects’ pre- and 1h post-transfusion PPP was measured. Results are reported as the change from pre-transfusion to 1h post-transfusion value for **(E)** thrombin generation peak (*p=0.0357), **(F)** lag time, **(G)** time to thrombin peak, and **(H)** start tail time. Data are shown as mean ± SEM and individual values for (**E-G**). n=9. The individual p-value is shown above.

**Supplemental Figure 4.**
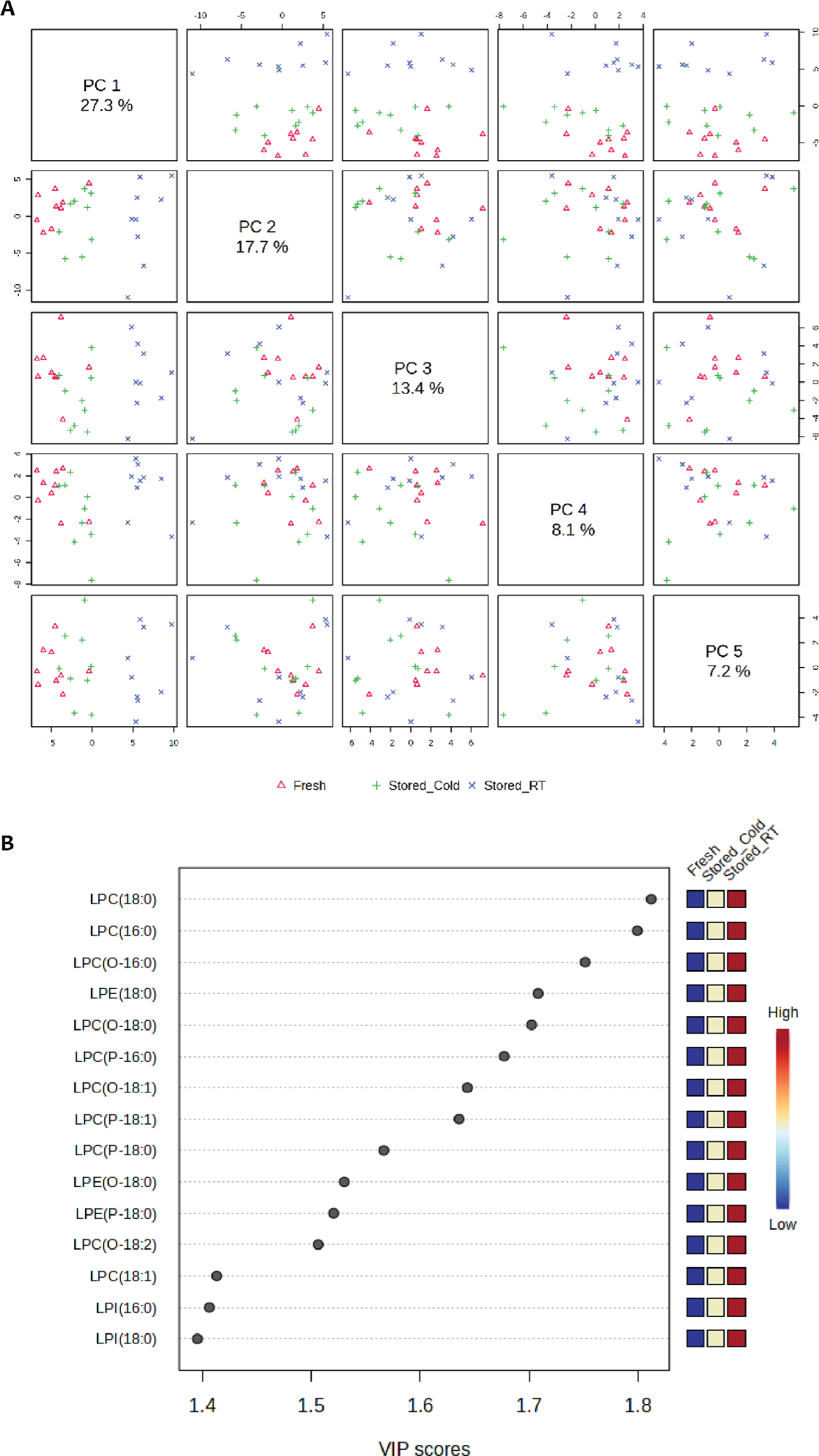
Lipidomic analyses of CSP and RSP. **(A)** Partial least square– discriminant analysis of platelet lipidomic phenotypes for Baseline, RSP and CSP. **(B)** The top 15 metabolites from the variable importance in projection plot.

**Supplemental Figure 5:**
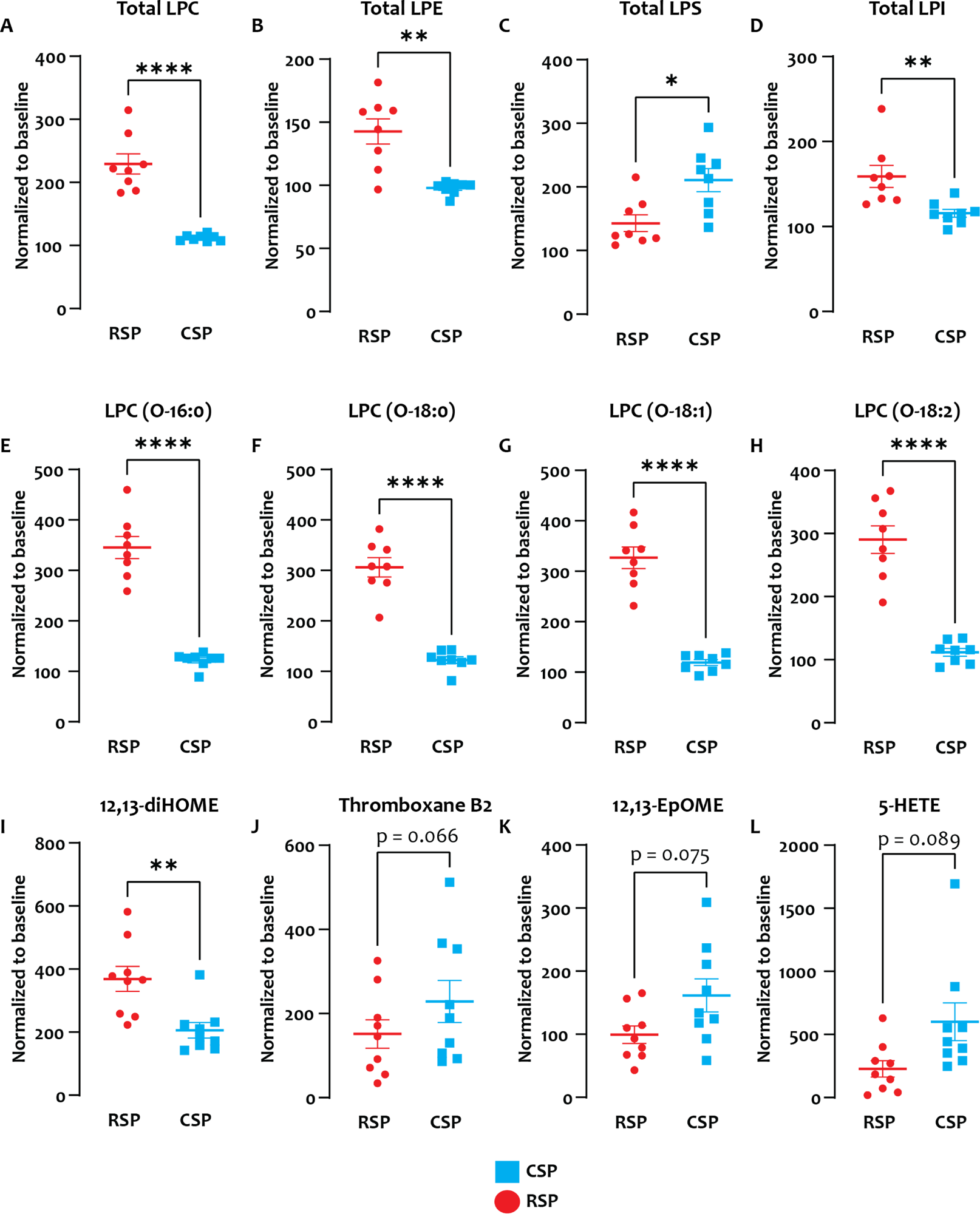
**A-D**: α = 0.05 with Holm-Sidak adjustment for multiple (4) comparisons: Total LPC p<0.0001, total LPE: p=0.002, Total LPS p=0.043, total LPI p=0.003. **E-L:** (Q=1% false discovery rate for multiple comparisons [89 comparisons]) p values: 16:0, 18:0, 18:1, 18:2: p<0.0001, 12,13-diHOME **p=0.0074

**Supplemental Figure 6:**
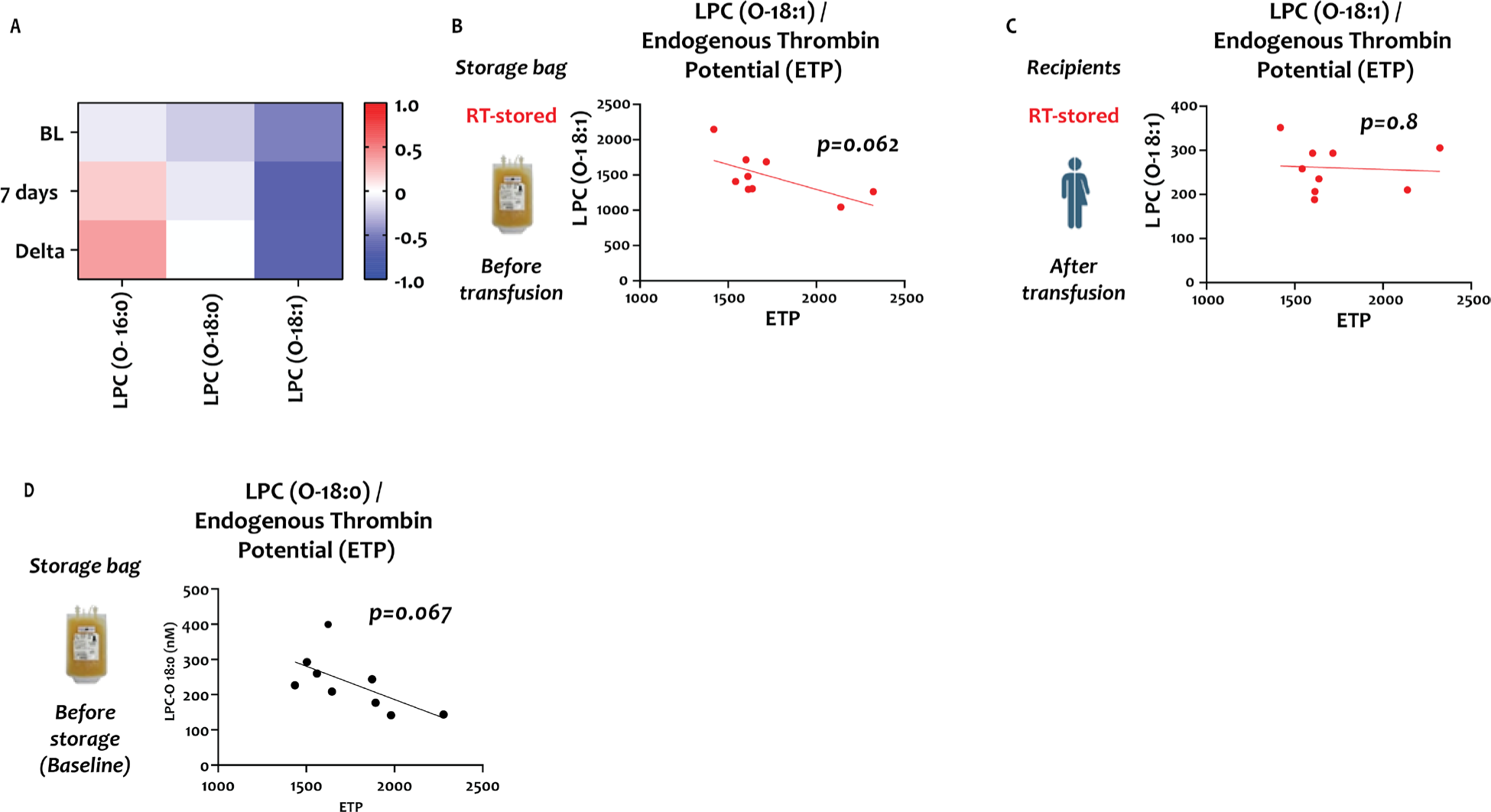
Correlation of lipid mediators and platelet function parameters. Left panel (**A**) shows r-value heat maps for baseline (BL), end of storage (7 days, respectively), and for the difference between BL and end of storage (Delta). Middle panel (**B**) shows the correlation between endogenous thrombin potential (ETP) and LPC(O-18:1) concentration. The right panel (**C**) shows the correlation between ETP and recipient LPC(O-18:1) concentration. Data are shown as heatmaps and linear regression analysis plots. N=8 in each group, paired data only, and individual p values are shown in each figure. (**D**) Correlation of LPC-O 18:0 at the *beginning of storage* and platelet function (ETP). Data are shown as linear regression analysis plot. N=8, paired data only, and individual p-values shown.

**Supplemental Table 1:** Demographics of study participants.

|  |  |
| --- | --- |
| Age, yr (IQR) | 33 (25,37) |
| Female sex, no (%) | 6 (60) |
| Height (cm) ± SD | 176 ± 11.5 |
| Weight (kg) ± SD | 88.4 ± 22.6 |
| BMI ± SD | 28 ± 4.7 |
| Washout (d) ± SD | 22 ± 8 |
| CSP first (%) | 5 (62) |

**Supplemental Table 2:**
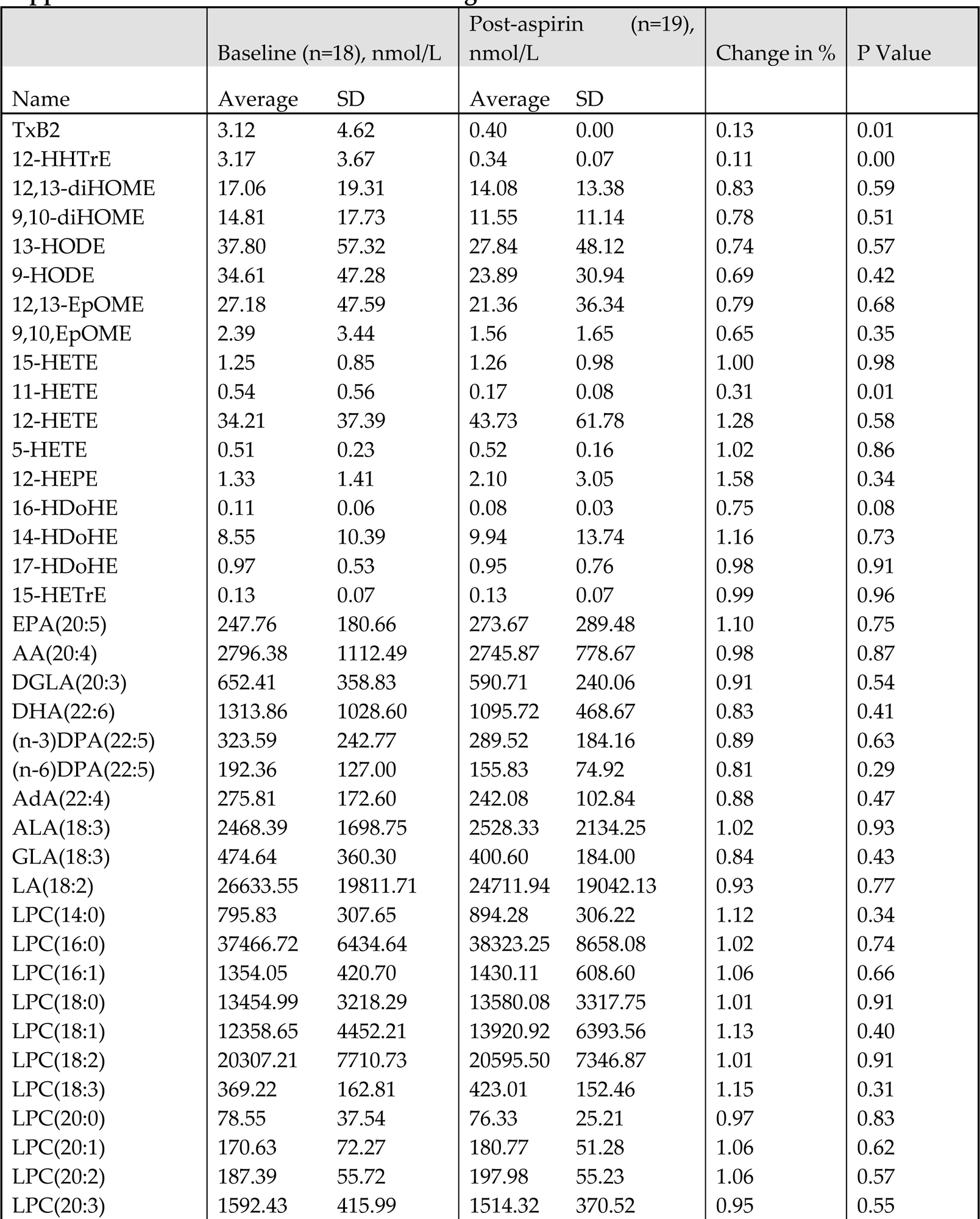
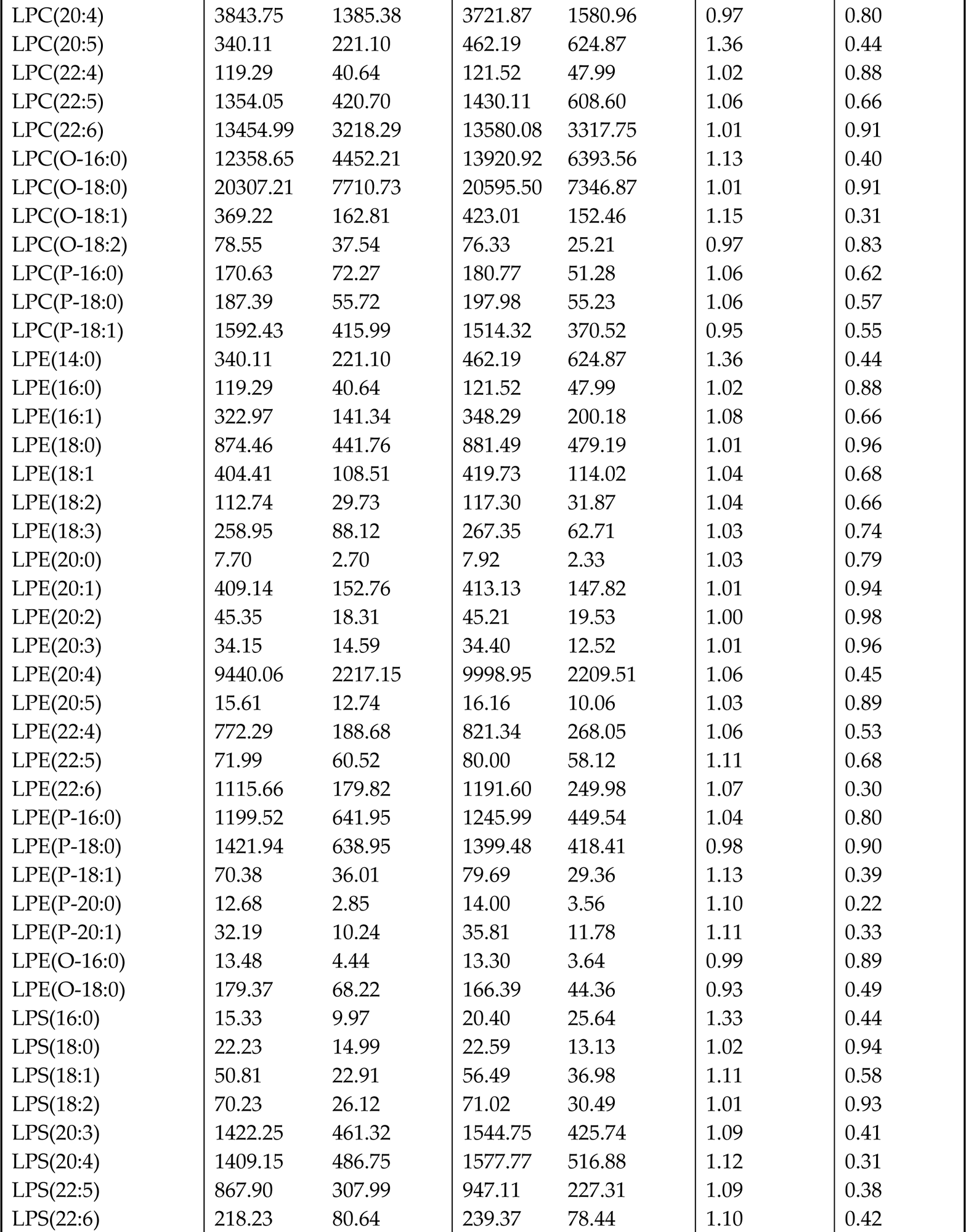
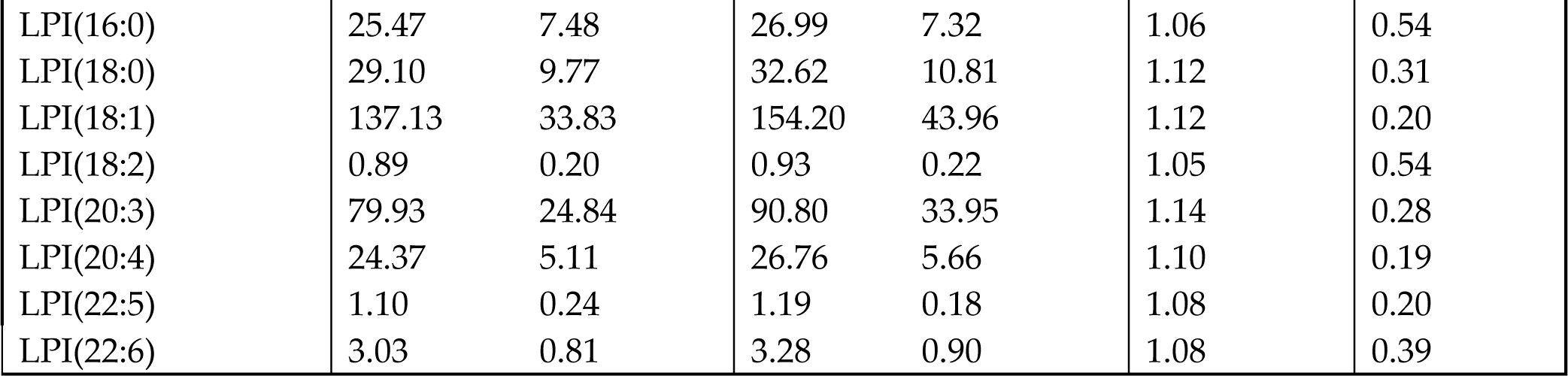
Concentration of targeted metabolites before and after ASA*.

|  | Baseline (n=18), nmol/L |  | Post-aspirin (n=19), nmol/L |  | Change in % | P Value |
| --- | --- | --- | --- | --- | --- | --- |
| Name | Average | SD | Average | SD |  |  |
| TxB2 | 3.12 | 4.62 | 0.40 | 0.00 | 0.13 | 0.01 |
| 12-HHTrE | 3.17 | 3.67 | 0.34 | 0.07 | 0.11 | 0.00 |
| 12,13-diHOME | 17.06 | 19.31 | 14.08 | 13.38 | 0.83 | 0.59 |
| 9,10-diHOME | 14.81 | 17.73 | 11.55 | 11.14 | 0.78 | 0.51 |
| 13-HODE | 37.80 | 57.32 | 27.84 | 48.12 | 0.74 | 0.57 |
| 9-HODE | 34.61 | 47.28 | 23.89 | 30.94 | 0.69 | 0.42 |
| 12,13-EpOME | 27.18 | 47.59 | 21.36 | 36.34 | 0.79 | 0.68 |
| 9,10-EpOME | 2.39 | 3.44 | 1.56 | 1.65 | 0.65 | 0.35 |
| 15-HETE | 1.25 | 0.85 | 1.26 | 0.98 | 1.00 | 0.98 |
| 11-HETE | 0.54 | 0.56 | 0.17 | 0.08 | 0.31 | 0.01 |
| 12-HETE | 34.21 | 37.39 | 43.73 | 61.78 | 1.28 | 0.58 |
| 5-HETE | 0.51 | 0.23 | 0.52 | 0.16 | 1.02 | 0.86 |
| 12-HEPE | 1.33 | 1.41 | 2.10 | 3.05 | 1.58 | 0.34 |
| 16-HDoHE | 0.11 | 0.06 | 0.08 | 0.03 | 0.75 | 0.08 |
| 14-HDoHE | 8.55 | 10.39 | 9.94 | 13.74 | 1.16 | 0.73 |
| 17-HDoHE | 0.97 | 0.53 | 0.95 | 0.76 | 0.98 | 0.91 |
| 15-HETrE | 0.13 | 0.07 | 0.13 | 0.07 | 0.99 | 0.96 |
| EPA(20:5) | 247.76 | 180.66 | 273.67 | 289.48 | 1.10 | 0.75 |
| AA(20:4) | 2796.38 | 1112.49 | 2745.87 | 778.67 | 0.98 | 0.87 |
| DGLA(20:3) | 652.41 | 358.83 | 590.71 | 240.06 | 0.91 | 0.54 |
| DHA(22:6) | 1313.86 | 1028.60 | 1095.72 | 468.67 | 0.83 | 0.41 |
| (n-3)DPA(22:5) | 323.59 | 242.77 | 289.52 | 184.16 | 0.89 | 0.63 |
| (n-6)DPA(22:5) | 192.36 | 127.00 | 155.83 | 74.92 | 0.81 | 0.29 |
| AdA(22:4) | 275.81 | 172.60 | 242.08 | 102.84 | 0.88 | 0.47 |
| ALA(18:3) | 2468.39 | 1698.75 | 2528.33 | 2134.25 | 1.02 | 0.93 |
| GLA(18:3) | 474.64 | 360.30 | 400.60 | 184.00 | 0.84 | 0.43 |
| LA(18:2) | 26633.55 | 19811.71 | 24711.94 | 19042.13 | 0.93 | 0.77 |
| LPC(14:0) | 795.83 | 307.65 | 894.28 | 306.22 | 1.12 | 0.34 |
| LPC(16:0) | 37466.72 | 6434.64 | 38323.25 | 8658.08 | 1.02 | 0.74 |
| LPC(16:1) | 1354.05 | 420.70 | 1430.11 | 608.60 | 1.06 | 0.66 |
| LPC(18:0) | 13454.99 | 3218.29 | 13580.08 | 3317.75 | 1.01 | 0.91 |
| LPC(18:1) | 12358.65 | 4452.21 | 13920.92 | 6393.56 | 1.13 | 0.40 |
| LPC(18:2) | 20307.21 | 7710.73 | 20595.50 | 7346.87 | 1.01 | 0.91 |
| LPC(18:3) | 369.22 | 162.81 | 423.01 | 152.46 | 1.15 | 0.31 |
| LPC(20:0) | 78.55 | 37.54 | 76.33 | 25.21 | 0.97 | 0.83 |
| LPC(20:1) | 170.63 | 72.27 | 180.77 | 51.28 | 1.06 | 0.62 |
| LPC(20:2) | 187.39 | 55.72 | 197.98 | 55.23 | 1.06 | 0.57 |
| LPC(20:3) | 1592.43 | 415.99 | 1514.32 | 370.52 | 0.95 | 0.55 |
| LPC(20:4) | 3843.75 | 1385.38 | 3721.87 | 1580.96 | 0.97 | 0.80 |
| LPC(20:5) | 340.11 | 221.10 | 462.19 | 624.87 | 1.36 | 0.44 |
| LPC(22:4) | 119.29 | 40.64 | 121.52 | 47.99 | 1.02 | 0.88 |
| LPC(22:5) | 1354.05 | 420.70 | 1430.11 | 608.60 | 1.06 | 0.66 |
| LPC(22:6) | 13454.99 | 3218.29 | 13580.08 | 3317.75 | 1.01 | 0.91 |
| LPC(O-16:0) | 12358.65 | 4452.21 | 13920.92 | 6393.56 | 1.13 | 0.40 |
| LPC(O-18:0) | 20307.21 | 7710.73 | 20595.50 | 7346.87 | 1.01 | 0.91 |
| LPC(O-18:1) | 369.22 | 162.81 | 423.01 | 152.46 | 1.15 | 0.31 |
| LPC(O-18:2) | 78.55 | 37.54 | 76.33 | 25.21 | 0.97 | 0.83 |
| LPC(P-16:0) | 170.63 | 72.27 | 180.77 | 51.28 | 1.06 | 0.62 |
| LPC(P-18:0) | 187.39 | 55.72 | 197.98 | 55.23 | 1.06 | 0.57 |
| LPC(P-18:1) | 1592.43 | 415.99 | 1514.32 | 370.52 | 0.95 | 0.55 |
| LPE(14:0) | 340.11 | 221.10 | 462.19 | 624.87 | 1.36 | 0.44 |
| LPE(16:0) | 119.29 | 40.64 | 121.52 | 47.99 | 1.02 | 0.88 |
| LPE(16:1) | 322.97 | 141.34 | 348.29 | 200.18 | 1.08 | 0.66 |
| LPE(18:0) | 874.46 | 441.76 | 881.49 | 479.19 | 1.01 | 0.96 |
| LPE(18:1) | 404.41 | 108.51 | 419.73 | 114.02 | 1.04 | 0.68 |
| LPE(18:2) | 112.74 | 29.73 | 117.30 | 31.87 | 1.04 | 0.66 |
| LPE(18:3) | 258.95 | 88.12 | 267.35 | 62.71 | 1.03 | 0.74 |
| LPE(20:0) | 7.70 | 2.70 | 7.92 | 2.33 | 1.03 | 0.79 |
| LPE(20:1) | 409.14 | 152.76 | 413.13 | 147.82 | 1.01 | 0.94 |
| LPE(20:2) | 45.35 | 18.31 | 45.21 | 19.53 | 1.00 | 0.98 |
| LPE(20:3) | 34.15 | 14.59 | 34.40 | 12.52 | 1.01 | 0.96 |
| LPE(20:4) | 9440.06 | 2217.15 | 9998.95 | 2209.51 | 1.06 | 0.45 |
| LPE(20:5) | 15.61 | 12.74 | 16.16 | 10.06 | 1.03 | 0.89 |
| LPE(22:4) | 772.29 | 188.68 | 821.34 | 268.05 | 1.06 | 0.53 |
| LPE(22:5) | 71.99 | 60.52 | 80.00 | 58.12 | 1.11 | 0.68 |
| LPE(22:6) | 1115.66 | 179.82 | 1191.60 | 249.98 | 1.07 | 0.30 |
| LPE(P-16:0) | 1199.52 | 641.95 | 1245.99 | 449.54 | 1.04 | 0.80 |
| LPE(P-18:0) | 1421.94 | 638.95 | 1399.48 | 418.41 | 0.98 | 0.90 |
| LPE(P-18:1) | 70.38 | 36.01 | 79.69 | 29.36 | 1.13 | 0.39 |
| LPE(P-20:0) | 12.68 | 2.85 | 14.00 | 3.56 | 1.10 | 0.22 |
| LPE(P-20:1) | 32.19 | 10.24 | 35.81 | 11.78 | 1.11 | 0.33 |
| LPE(O-16:0) | 13.48 | 4.44 | 13.30 | 3.64 | 0.99 | 0.89 |
| LPE(O-18:0) | 179.37 | 68.22 | 166.39 | 44.36 | 0.93 | 0.49 |
| LPS(16:0) | 15.33 | 9.97 | 20.40 | 25.64 | 1.33 | 0.44 |
| LPS(18:0) | 22.23 | 14.99 | 22.59 | 13.13 | 1.02 | 0.94 |
| LPS(18:1) | 50.81 | 22.91 | 56.49 | 36.98 | 1.11 | 0.58 |
| LPS(18:2) | 70.23 | 26.12 | 71.02 | 30.49 | 1.01 | 0.93 |
| LPS(20:3) | 1422.25 | 461.32 | 1544.75 | 425.74 | 1.09 | 0.41 |
| LPS(20:4) | 1409.15 | 486.75 | 1577.77 | 516.88 | 1.12 | 0.31 |
| LPS(22:5) | 867.90 | 307.99 | 947.11 | 227.31 | 1.09 | 0.38 |
| LPS(22:6) | 218.23 | 80.64 | 239.37 | 78.44 | 1.10 | 0.42 |
| LPI(16:0) | 25.47 | 7.48 | 26.99 | 7.32 | 1.06 | 0.54 |
| LPI(18:0) | 29.10 | 9.77 | 32.62 | 10.81 | 1.12 | 0.31 |
| LPI(18:1) | 137.13 | 33.83 | 154.20 | 43.96 | 1.12 | 0.20 |
| LPI(18:2) | 0.89 | 0.20 | 0.93 | 0.22 | 1.05 | 0.54 |
| LPI(20:3) | 79.93 | 24.84 | 90.80 | 33.95 | 1.14 | 0.28 |
| LPI(20:4) | 24.37 | 5.11 | 26.76 | 5.66 | 1.10 | 0.19 |
| LPI(22:5) | 1.10 | 0.24 | 1.19 | 0.18 | 1.08 | 0.20 |
| LPI(22:6) | 3.03 | 0.81 | 3.28 | 0.90 | 1.08 | 0.39 |

