## Supplemental for "Platelet dysfunction reversal with cold-stored vs. room temperature-stored platelet transfusions"

**METHODS**

*Reagents, Materials,* Platelet agonists ADP, arachidonic acid, and fibrillar collagen type I (all from Chronolog, Havertown, PA). Samples for VerifyNOW were sent to the University of Washington clinical laboratory at UW Harborview (Seattle WA). ASA was purchased from the University of Washington Pharmacy. Platelets antibodies were purchased from BD Pharmingen (Franklin Lakes, New Jersey, USA). Platelet counts were measured with a commercially available hemocytometer (ABX Micros, Horiba, Irvine, CA, USA).

*Unit characteristics and safety assessment.*

All platelet units passed quality control assessments after storage, except for one RSP unit that had to be discarded because of low pH. Transfused platelets were tolerated well by all recipients. One subject developed a headache, which was considered unrelated to transfusion. The absolute number of platelets transfused was significantly lower in the CSP group. However, the transfused platelets as a percentage of all circulating platelets did not differ between both groups (Supplemental Figure 3 A, B).

*Thrombin generation assay*

Platelets were obtained from transfusion recipients and sampled from storage bags. When taken from the storage bag, platelets were diluted from storage concentration with separately-stored, autologous fresh frozen plasma (stored at -80°C on the day of collection) from the same donor to dilute platelets to a target concentration of $1x10^5/\mu L$. The diluted platelets were tested in a commercially available fluorogenic Stago Thrombinoscope assay (Stago, Parsippany, NJ, USA). Platelet-rich plasma samples were stimulated with the PRP reagent (mainly tissue factor and minimal phospholipids), and when we tested the platelet-poor plasma (PPP) samples for thrombin generation we used the reagent containing only phospholipids (Microparticle reagent, both by Stago, Parsippany, NJ, USA) to stimulate thrombin generation. We also used a PPP reagent (TF and phospholipids for PPP samples, but results did not differ between microparticle (MP) and PPP reagents (data not shown).

*Light transmission aggregometry*

Platelets were isolated from whole blood as platelet-rich plasma by one-step soft centrifugation and adjusted to 300k/μL with autologous platelet-poor plasma (PPP). We performed the experiment at 37°C under stirring conditions (1200 rpm). After the addition of agonists, light transmission was recorded over 7 min on a Chrono-log 4-channel optical aggregation system (Chrono-log, Havertown, PA).

*Platelet microparticle characterization*

Samples for flow cytometry were tested with a CytoFLEX V4-B4-R3 Flow Cytometer (Brea, CA, USA). Beads were purchased (Gigamix Labling Kit (Megamix-Plus FSC reagent (0.1 μm, 0.3 μm, 0.5 μm and 0.9 μm.) plus Megamix-Plus SSC reagent (0.16 μm,

0.20 μm, 0.24 μm and 0.5 μm.) from Biocytex (Marseille, France). Microparticles (MP) were identified by size and defined as large PMP (0.5 μm-eq. to 1μm) and small (0.1 μm to 0.5 μm-eq.) with the help of nanobeads, as outlined in Figure 3 A-C. Platelet MP (PMP) were identified in the MP gates and defined as CD41a positive events and further characterized as phosphatidyl serine positive events with FITC-labeled lactadherin (Haematologic Technologies Prolytix [HTI] Essex Junction, Vermont, USA) and as P-selectin positive with anti-CD62P-PE (both BD Pharmingen, NJ, USA) (Figure 3 D-F). Samples were acquired at 10μL/min, and a few modifications were performed: we switched the 405 vs 450 filters to help with size differentiation, and we also used the PB450 as the SSC channel for the gating strategy. We also increased the threshold based on PB450 up to 100nm to reduce the noise. The cytometer was thoroughly cleaned with deionized water between every run. A washing solution was used at the beginning and end of every plate run. All solutions and reagents were microfiltered to reduce noise.

*Analysis of free PUFAs, oxylipins, and lysoglycerophospholipids*

As described previously, free PUFAs, oxylipins, and lysoglycerophospholipids in platelets were analyzed by liquid chromatography-tandem mass spectrometry with multiple reaction monitoring (LC-MS/MS-MRM). Briefly, analytes were extracted by 80% methanol (vol/vol) with an internal standard mixture containing deuterium labeled analogs (Cayman Chemical) and 17:1 lysophospholipids (Avanti Polar Lipids). Analytes were separated on a C18 column (Acquity HSS T3, 2.1 × 100 mm, 1.8 μm, Waters). The mobile phase was composed of (A) water/acetonitrile (95/5, vol/vol) with 5 mmol/L ammonium acetate and (B) 2-propanol/acetonitrile/water (50/45/5, vol/vol/vol) with 5 mmol/L ammonium acetate. Analytes were detected using multiple reaction monitoring (MRM) in the negative ion mode and were quantified by peak area relative to each corresponding internal standard. Data were collected and processed using computer software (Analyst Version 1.6.2, MultiQuant Version 2.1.1, AB Sciex).
